# Trajectories of and spatial variations in HPV vaccine discussions on Weibo, 2018-2023: a deep learning analysis

**DOI:** 10.1101/2023.12.07.23299667

**Authors:** You Wang, Haoyun Yang, Zhijun Ding, Xinyu Zhou, Yingchen Zhou, Liyan Ma, Zhiyuan Hou

## Abstract

**Research in context:** *Evidence before this study:* We first searched PubMed for articles published until November 2023 with the keywords “(“HPV”) AND (“Vaccine” or “Vaccination”) AND (“Social Media”)”. We identified about 390 studies, most of which were discussions on the potentials or feasibility of social media in HPV vaccination advocacy or research, or manual coding-driven analyses on text (eg., tweets) about HPV vaccines emerged on social media platforms. When we added keyword “Machine Learning”, we identified only 12 studies, with several of them using AI-driven approach, such as deep learning, machine learning, and natural language process, to analyze extensive text data about public perceptions of HPV vaccination and perform monitor on social media platforms, X (Twitter) and Reddit. All these studies are from English-language social media platforms in developed countries. No study to date has monitored public perceptions of HPV vaccination on social media platforms from the developing countries including China.

*Added value of this study:* This is the first deep-learning study monitoring public perceptions of HPV vaccination expressed on Chinese social media platforms (Weibo in our case), revealing key temporal and geographic variations. We found a sustained high level of positive attitude towards HPV vaccination and exposure to social norms facilitating HPV vaccination among Weibo users, with a lower national prevalence of negative attitude, perceived barriers to accepting vaccination, misinformation about HPV or HPV vaccination, indicating the achievement of relevant health communication. High prevalence practical barriers to HPV vaccination expressed on Weibo was associated with relatively insufficient of HPV vaccine accessibility in China, suggesting the health systems should prioritize on addressing issues about vaccine supply. Lower positive perception of HPV vaccination among male users, higher vaccine hesitancy towards 2-valent vaccine, and provincial-level spatial cluster of higher negative attitude towards HPV vaccination indicate that tailored strategies need to be formed targeting specific population, areas, and vaccine type. Our monitor practice on public perceptions of HPV vaccine from Weibo shows the feasibility of realizing public health surveillance potential of social media listening in Chinese context. Leveraging recent advances in deep learning, our approach could be a cost-effective supplement to existing surveillance techniques.

*Implications of all the available evidence:* This national surveillance study highlights the value of deep learning-driven social media listening as a convenient and effective approach for identifying emerging trends in public perceptions of HPV vaccination to inform interventions. As a supplement to existing public health surveillance techniques, it is particularly helpful to inform tailored and timely strategies in health communication and resource allocation at multiple levels. Key stakeholders and officials should maintain focus on health education highlighting the risks and consequences of HPV infections, and benefits and safety of all types of HPV vaccines; health systems should aim to resolve issues of vaccine accessibility. A proposed research area is the further development of deep learning models to monitor public perceptions of vaccines and analyzing misinformation about and barriers to HPV vaccination expressed on Chinese social media platforms.

**Background:** HPV vaccination rate is low in China. Understanding the multidimensional barriers and impetuses perceived by individuals to vaccination is essential. We aim to assess the public perceptions, perceived barriers, and facilitators towards HPV vaccination expressed on Chinese social media platform Weibo.

**Methods:** We collected Weibo posts regarding HPV vaccines between 2018 to 2023. We annotated 6,600 posts manually according to behavior change theories, and subsequently fine-tuned deep learning models to annotate all posts collected. Based on the annotated results of deep learning models, temporal and geographic analyses were conducted for public attitudes towards HPV vaccination and its determinants.

**Findings:** Totally 1,972,495 Weibo posts were identified as relevant to HPV vaccines. Deep learning models reached predictive accuracy of 0.78 to 0.96 in classifying posts. During 2018 to 2023, 1,314,510 (66.6%) posts were classified as positive attitudes. And 224,130 posts (11.4%) were classified as misinformation, 328,442 posts (16.7%) as perceived barriers to accepting vaccines, and 580,590 posts (29.4%) as practical barriers to vaccination. The prevalence of positive attitude increased from 15.8% in March 2018 to 79.1% in mid-2023 (p < 0.001), and misinformation declined from 36.6% in mid-2018 to 10.7% in mid-2023 (P < .001). Central regions exhibited higher prevalence of positive attitudes and social norms, whereas Shanghai, Beijing megacities and northeastern regions showed higher prevalence of negative attitudes and misinformation. Positive attitudes were significantly lower for 2-valent vaccines (65.7%), than 4-valent or 9-valent vaccines (79.6% and 74.1%).

**Interpretation:** Social media listening represents a promising surveillance approach for monitoring public perceptions on health issues and can enable the development of health communication strategies.

## Introduction

As a significant public health challenge, human papillomavirus (HPV) infections contribute to an annual occurrence of approximately 108,000 cervical cancer cases in China, and lead to incidence in anogenital and head and neck cancers.^1,2^ HPV vaccine has been proved to effectively prevent HPV infections, and included in the national immunization programme in 136 countries.^3^ However, it has not been included in China’s national immunization programme, with only 2.24% of the vaccine-eligible age population vaccinated.^4^ In China, stigma against HPV, rooted in conservative sexual norms, impedes open discussion on sexual health. This cultural backdrop leads to limited awareness and hesitancy towards HPV vaccination, often confined by misconceptions about its necessity, influenced by traditional views on sexual activity and health education.^5^ As social media becomes popular, the young generation posts massive discussions about HPV vaccine on social media, which may drive changes in sexual norms and public attitudes towards the HPV vaccine.^6^ In addition, few cities in China started to introduce HPV vaccines into local immunization programme since 2022, promoting public attention to HPV vaccine.^7^ Therefore, it is imperative to investigate discussions on social media and its role in HPV vaccination. It will help to map the dynamics of public perceptions, perceived barriers, and facilitators towards HPV vaccination in China.

Behavior change theories provide frameworks to understand the dynamics of attitudes and behaviors towards HPV vaccination in China. Among them, the Health Belief Model (HBM) and Theory of Planned Behavior (TPB) stand theoretically and empirically validated,^8,9^ serving as the foundational frameworks. Adapted from HBM and TPB, the Increasing Vaccination Model has been developed for the vaccination field and adopted by the World Health Organization.^10–12^ This model centers on vaccination motivation and vaccine hesitancy, which are predicted by internal health beliefs and external information environments, and it also includes practical issues when conducting actual vaccination behaviors with positive motivation.^12^

Understanding the public’s perceptions is essential to develop tailored education strategies and promote HPV vaccination. Social media listening can play a crucial role in assessing public perceptions and assisting policy-makers as a supplementary to traditional methods.^13,14^ With the incorporation of advanced machine learning techniques, social media data transcends simple textual or visual information and constructs a nuanced narrative that reveals public attitudes and actions.^15^ Social media listening allows to scrutinize the ever-changing dynamics of public attitudes towards health issues and reveal how social environments reshape public attitudes in an economically efficient and expeditious way.^16^ As a subset of machine learning techniques, deep learning (DL) with fine-tuning by gold-standard corpus (i.e., manual pre-annotated dataset) has exhibited remarkable performance advantages in analyzing social media data.^17,18^

Social media listening has been widely practiced during the COVID-19 pandemic.^19^ It was commonly applied to investigate the impacts of pandemic containment policies on public sentiment,^20^ and track social sentiments, topics, and the spread of misinformation regarding COVID-19 vaccines.^21^ Few studies has employed social media platforms to assess public perceptions towards HPV vaccination,^22,23^ but mainly focused on English-language platforms such as X and YouTube.^24^ There is limited knowledge on the perception of HPV vaccine among social media users in China.

Our study aimed to perform a deep learning analysis of HPV vaccine-related discussions posted on Weibo, the Chinese counterpart of X in the social media landscape, from early 2018 to mid-2023. We fine-tuned and deployed DL techniques to annotate millions of Weibo posts regarding HPV vaccines according to behavior change theories. Based on the DL annotation results, we assessed the temporal and spatial trends in public perceptions, perceived barriers, and facilitators towards HPV vaccination in China.

## Methods

An overview of the study design is presented in Figure 1. Methods in details are shown in supplementary materials.

**Figure 1.**
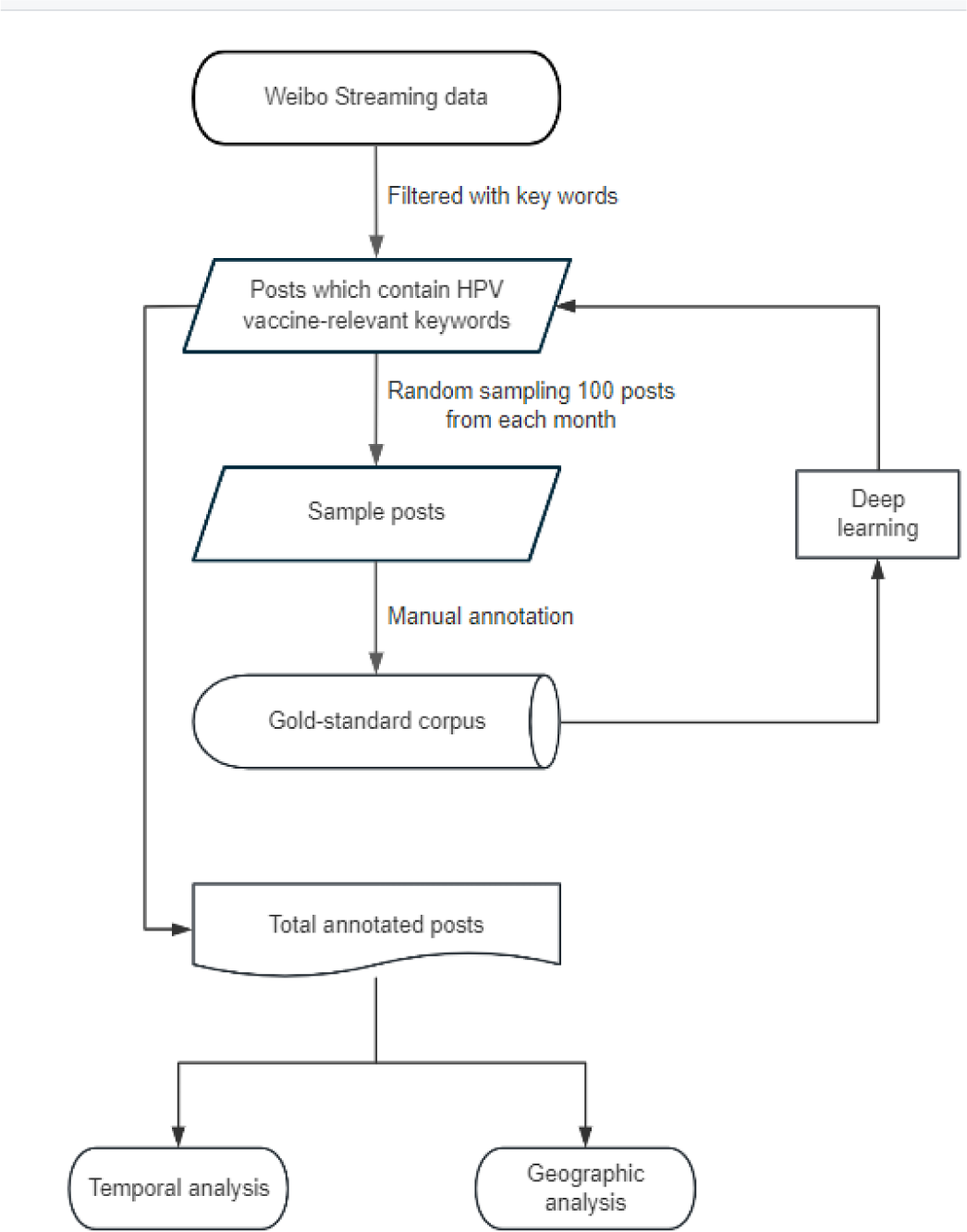
Overview of Study Design

### Data Collection

Weibo was leveraged as our data source due to its substantial user base in China and the accessibility of well-established streaming data acquisition channels. We conducted a data screening process using a predefined set of keywords to capture posts through Weibo application programming interface (API). Since there are 2-valent (2v), 4-valent (4v), and 9-valent (9v) vaccines available in China, the keywords included “HPV vaccine” (“••癌疫苗”, “HPV 疫苗”), “vaccinate against HPV” (“打 HPV”), “9-valent” (“9 价”), “nine-valent” (“九价”), “4-valent” (“4 价”), “four-valent” (“四价”), “2-valent” (“2 价”), “two-valent” (“二价”). The resulting dataset comprises a total of 4,154,274 posts, spanned from January 1, 2018, to June 30, 2023. We also collected self-reported gender and province-level location data for each posting account. Some posts may have no location or gender data since Weibo users have the option not to disclose their locations or gender profile.

### Framework and Manual Annotation

Adapted from the Increasing Vaccination Model, HBM, and TPB,^10,25^ a conceptual framework (Figure 2) was developed to guide our annotation on Weibo posts. In this conceptual framework, health beliefs and information environments influence the attitudes towards vaccination, and practical barriers to vaccination moderates the process from attitudes (ie., positive, neutral, negative) to behaviors. As internal factors of HPV vaccination, health beliefs include three constructs – perceived disease risks, perceived benefits of vaccines, and perceived barriers to accepting vaccines. As external factors of HPV vaccination, information environment includes two constructs - misinformation and social norms. Among these framework constructs, perceived disease risks, perceived benefits of vaccines, and social norms can be seen as facilitators towards HPV vaccination, whereas perceived barriers to accepting vaccines, misinformation, practical barriers to vaccination can be seen as barriers to HPV vaccination. Definitions of framework constructs and corresponding raw examples of posts are presented in the supplementary material (eTable 1).

**Figure 2.**
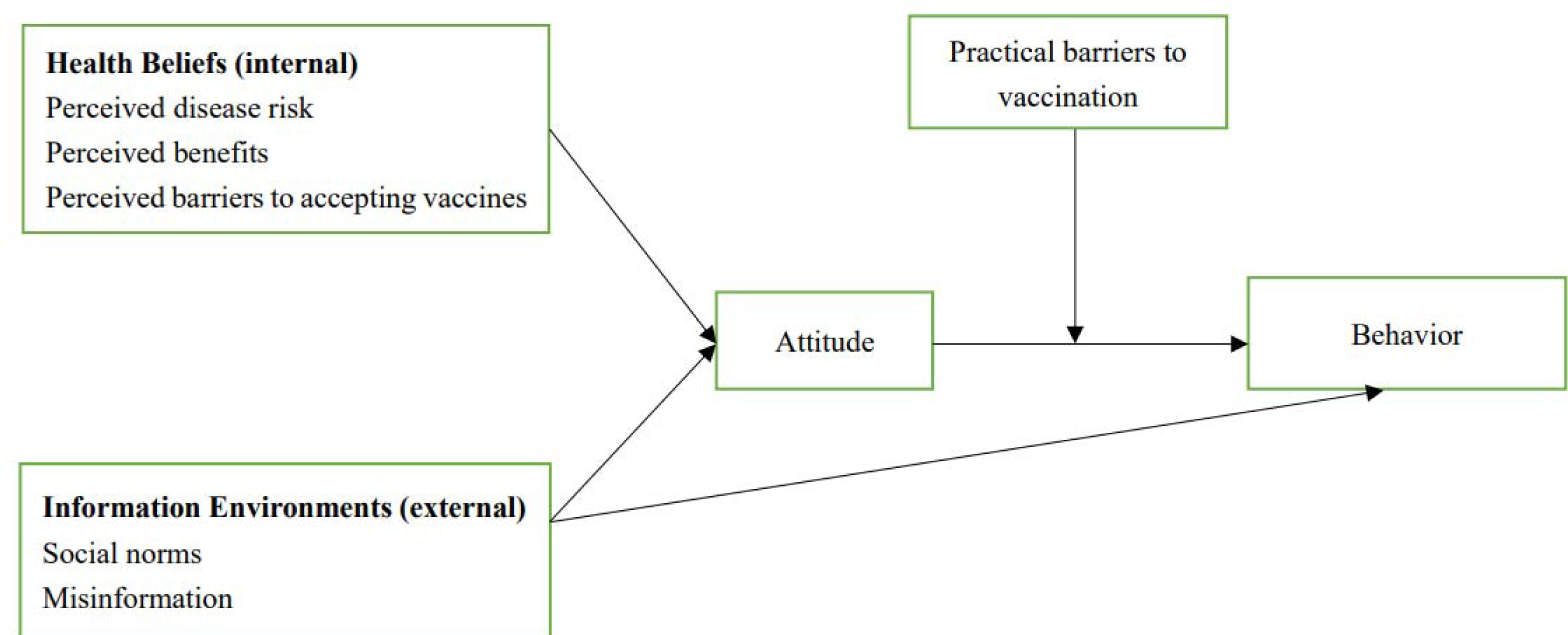
Conceptual framework of HPV vaccination NOTE: Health beliefs indicate constructs of individual’s internal beliefs about HPV and HPV vaccine, including perceived disease risk of HPV, perceived benefits of vaccines and perceived barriers to accepting vaccines; environments indicate constructs of individual’s exposure to environmental barriers or facilitators regarding HPV vaccination, including social norms facilitating HPV vaccination, misinformation about HPV or HPV vaccines, and practical barriers to vaccination. Locally estimated scatterplot smoothing (LOESS) is employed to reveal the temporal trends of prevalence and remove random noise components on monthly raw data.

Manually annotated gold-standard corpus of HPV vaccine-related Weibo posts is essential to fine-tune DL algorithms for analyzing HPV vaccine-related posts. We performed random extraction of 100 posts per month within the observation period, resulting in a subset of 6,600 posts in total for manual annotation. Two annotators independently annotated each post according to its relevance to the constructs in the conceptual framework. Firstly, annotators coded each post’s overall attitude toward the HPV vaccine as positive, negative, or neutral. Afterwards, each post was annotated according to its relevance to the remaining six constructs as factors of HPV vaccination. A post could be relevant to one or more constructs or to none. Cohen’s κ interrater agreement between coders was 0.933 for attitude annotation, and ranged from 0.846 to 0.980 for each non-attitude construct (eTable 2, in the supplementary material). This meticulously curated gold-standard corpus of 6,600 manually annotated posts served as the foundation for fine-tuning and evaluating DL algorithms.

### Fine-tuning Deep Learning Models for Classification

We applied the SetFit (Sentence Transformer Fine-tuning) approach to auto-annotate the total sample of Weibo posts, which is a few-shot classification approach based on the Sentence Transformer models.^26^ Sentence Transformer commonly adds a (mean) pooling layer to pre-trained encoder-only models like vanilla BERT (Bidirectional Embedding Representation Transformer) and were trained using datasets regarding sentence relationship to generate sentence-level embeddings (i.e., high-dimensional vectors).^27^ It can be coupled with a classification head downstream to accomplish classification tasks, enhancing the training efficiency.^27^ The SetFit model comprises two training phases: the first involves fine-tuning Sentence Transformer, and the second involves training the classification head. This approach can achieve classification efficacy through few-shot training without prompts and yield high few-shot performance.

We adopted a two-stage hierarchical classification strategy to fine-tune multiple DL classification models using 6,600 manually annotated posts as gold-standard corpus. The first stage is a fine-tuned DL classification model to distinguish whether a post is related to the HPV vaccine or not, and the second stage includes seven DL models for labeling HPV vaccine-relevant posts to each of all seven constructs in the annotation framework. For each DL model, we randomly selected 80% of the manually annotated data as the training set and reserved 20% as the test set, and all the DL models achieved an accuracy over 0.78 (Table 1). See details in supplementary materials.

**Table 1.** Framework constructs and predictive performance of SetFit models in Weibo posts.

| Framework construct | Predictive performance of SetFit models |  |  |  |
| --- | --- | --- | --- | --- |
|  | Accuracy | Precision | Recall | F-1 score |
| **Irrelevant** | 0.9572<br>(0.9426-0.9718) | 0.9925<br>(0.9863-0.9986) | 0.9572<br>(0.9426-0.9718) | 0.9747<br>(0.9665-0.9829) |
| **Attitude** | 0.7829<br>(0.7132-0.8527) | 0.7600<br>(0.6774-0.8427) | 0.7490<br>(0.6677-0.8303) | 0.7508<br>(0.6700-0.8316) |
| **Health Beliefs** |  |  |  |  |
| Perceived disease risk | 0.8827<br>(0.8148-0.9506) | 0.6310<br>(0.4286-0.8333) | 0.9300<br>(0.7778-1.) | 0.7325<br>(0.5714-0.8936) |
| Perceived benefits | 0.8567<br>(0.8000-0.9133) | 0.7830<br>(0.6591-0.9070) | 0.7353<br>(0.6078-0.8627) | 0.7545<br>(0.6575-0.8515) |
| Perceived barriers to accepting vaccines | 0.8645<br>(0.8037-0.9252) | 0.7630<br>(0.6286-0.8974) | 0.8492<br>(0.7317-0.9667) | 0.7987<br>(0.700-0.8974) |
| **Environments** |  |  |  |  |
| Social norms | 0.8343<br>(0.7952-0.8735) | 0.7750<br>(0.6970-0.8529) | 0.7425<br>(0.6636-0.8214) | 0.7568<br>(0.6946-0.8190) |
| Misinformation | 0.8862<br>(0.8293-0.9431) | 0.8152<br>(0.6875-0.9429) | 0.7698<br>(0.6333-0.9063) | 0.7867<br>(0.6792-0.8941) |
| Practical barriers to vaccination | 0.8592<br>(0.8338-0.8846) | 0.7893<br>(0.7368-0.8417) | 0.7760<br>(0.7236-0.8283) | 0.7819<br>(0.7414-0.8224) |

Note: Mean (95% CI) are shown.

### Statistical Analysis

We performed statistical analyses on the DL-generated prevalence for all framework constructs using R (Version 4.3.1), including both temporal and geographic analyses. All tests were two-tailed and the level of statistical significance was established at P < .05.

Firstly, we performed the temporal analysis by calculating the monthly prevalence of posts relating to each framework construct. We employed locally estimated scatterplot smoothing (LOESS) to reveal the temporal trends of prevalence and remove random noise components.^28^ Time-series analyses were used to identify whether and when these constructs exhibited statistically significant increases or decreases in trends.

Secondly, we performed geographic analysis at provincial level, focusing on spatial distribution of the prevalence of each framework construct. We used self-reported location from user profiles to identify geographic location of each post.

We also compare the prevalence of framework constructs by different valent types of HPV vaccines and users’ gender. We utilized keyword-based retrieval to classify relevant posts into the three valent types, each retaining only posts exclusively mentioning 2v, 4v, or 9v vaccines. We also distinguished relevant posts by gender self-reported in users’ profiles.

### Role of funding

The funders have no role in study design, data analysis and interpretation of data, the writing of the manuscript, or the decision to submit the paper for publication.

## Results

Among all 4,154,274 Weibo posts from keyword-based screening, 1,972,495 posts from 1,263,324 unique users were identified as relevant to HPV vaccines and based on mainland China, which were used for our statistical analyses. Among these, 1,217,389 posts (61.7%) from 803,238 users (63.6%) were identified with provincial locations. The fine-tuned SetFit models demonstrated robust predictive performance and consistent accuracy (Table 1), ranging from 0.7829 (95% CI, 0.7132-0.8527) to 0.9572 (95% CI, 0.9426-0.9718), and achieved mean F-1 scores ranging from 0.7325 (95% CI, 0.5714-0.8936) to 0.9747 (95% CI, 0.9665-0.9829).

### Overall Status of the Framework Constructs

During the study period from January 2018 to June 2023, 1,314,510 (66.6%), 120,717 (6.1%), and 537,268 (27.2%) posts were classified as positive, negative, and neutral attitudes towards HPV vaccines, respectively. Regarding internal health beliefs, 233,130 posts (11.8%) were classified as perceived disease risks, 245,622 posts (12.5%) as perceived benefits of vaccines, and 328,442 posts (16.7%) as perceived barriers to accepting vaccines. Regarding exposures to external information environments and practical issues, 432,754 posts (21.9%) were classified as social norms, 224,130 posts (11.4%) as misinformation, and 580,590 posts (29.4%) as practical barriers to vaccination.

### Temporal Trends of the Framework Constructs

The temporal trends with smoothing techniques, for the number of relevant posts and the prevalence of each framework construct, are illustrated in Figure 3, with raw data in the supplementary material (eFigure 1). Over the study period, we observed a noteworthy increase in the number of HPV vaccine-related posts after 2020, with the peak in the first half of 2022. For attitudes towards the HPV vaccine, the prevalence of positive attitude had notably increased since March 2018, from 15.8% to 79.1% (p < 0.001). Concurrently, the prevalence of negative attitude displayed a declining trend (p < 0.05), especially after its peak in August 2019 (20.3%) to the end of our observation period in June 2023 (5.5%; P < 0.001). Obviously, the prevalence of positive attitude far outweighed the prevalence of negative attitude.

**Figure 3.**
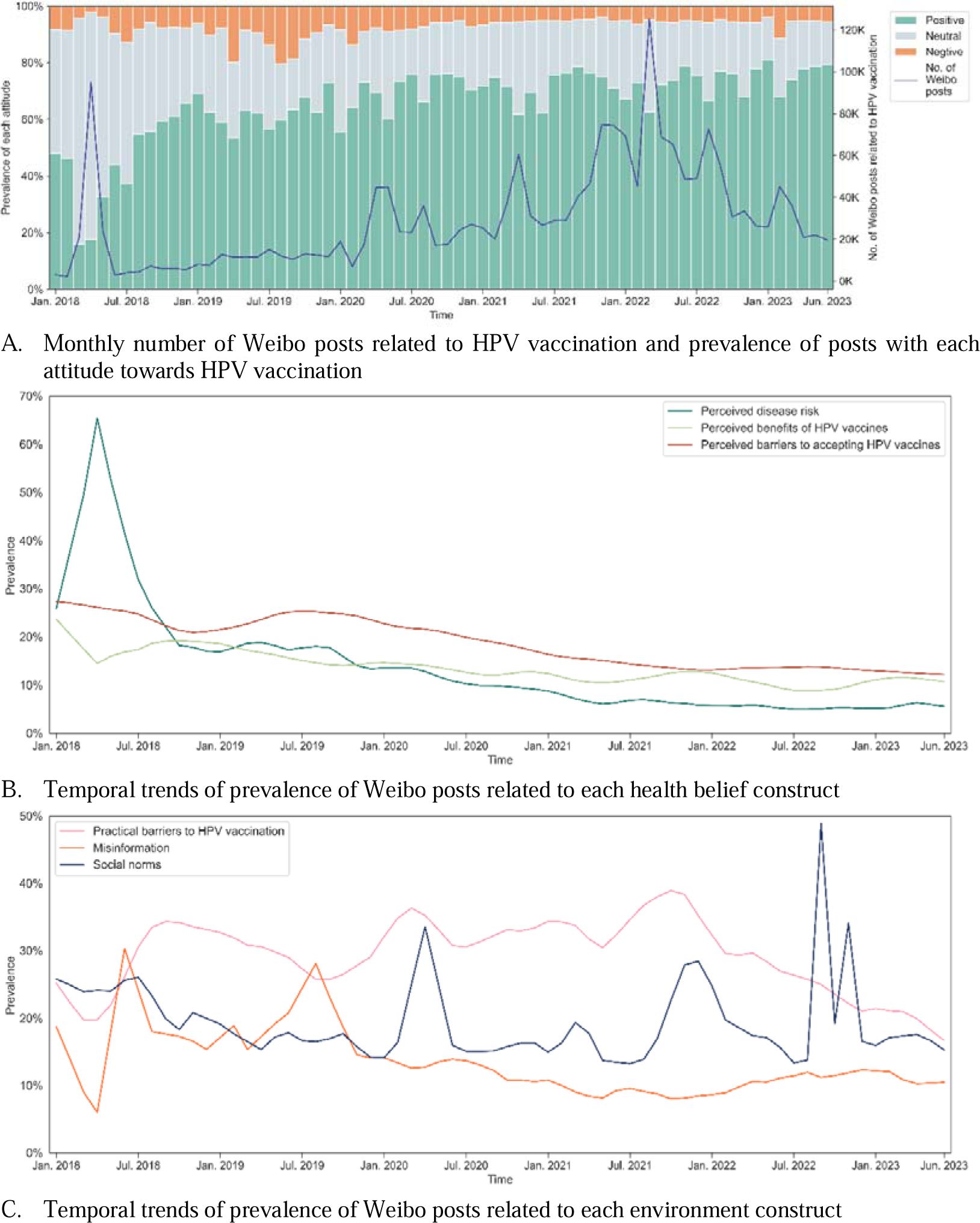
Temporal trends of the number of Weibo posts and prevalence of posts related to each framework construct of HPV vaccination NOTE: Insufficient data indicates regions excluded from geographic analyses: 1) Hong Kong, Macau, and Taiwan because of different HPV vaccination policy from mainland China; 2) Qinghai, Tibet, and Ningxia (3651, 3709, and 3749 posts) because of too small sample size.

Among the constructs of internal health beliefs, the prevalence of perceived disease risks declined, from its peak in April 2018 (65.4%) to June 2023 (where our observation ended; 4.5%; P < 0.001). Similarly, the prevalence of perceived benefits of vaccines declined significantly from July 2018 (30.4%) to June 2023 (9.9%; P < 0.001). The prevalence of perceived barriers to accepting vaccines had been staying high and stable since July 2018 (34.6%) until February 2020 (36.9%; P = 0.84), and then decreased extending to June 2023 (13.1%; P < 0.001). While constantly decreasing, the prevalence of perceived benefits was significantly lower than that of perceived barriers (mean difference, −5%; P < 0.001).

Among the constructs of external environments, the overall prevalence of social norms stabilized (P = 0.87) with a few outliers (the highest point in September 2022, 48.9%). It only decreased slightly from early 2018 to the lowest in February 2021 (12.7%; P = 0.006), and remained consistent thereafter (P = 0.94). Misinformation decreased from the middle of 2018, with the peak in July 2018 (36.6%), down to the middle of 2023 (10.7%; P < 0.001). Noteworthy is that all time points with the higher prevalence of misinformation were located during 2018 and 2019 (June 2018, 30.3%; July 2018, 36.6%; April 2019, 32.4%; July 2019, 30.6%; August 2019, 28.1%). The prevalence of practical barriers to vaccination increased from early 2018 to its peak in August 2020 (46.8%; P < 0.05), and then decreased (June 2023, 17.2%; P < 0.001). Over the observation period, the prevalence of practical barriers sustained significantly higher than social norms and misinformation. While the prevalence of social norms was generally higher than misinformation, their relationship varied temporally: from January 2018 to March 2020, no significant difference was observed; however, from March 2020 to June 2023, the prevalence of social norms was significantly greater (mean difference 9.5%; P < 0.001).

### Geographical Variations of the Framework Constructs

The inter-province variations, for the number of relevant posts and the prevalence of attitudes, are depicted in Figure 4, and for constructs of health beliefs and environments in the supplementary material (eFigure 2). Monthly numbers of relevant posts by province are depicted in the supplementary material (eFigure 3). The number of relevant posts revealing a significant regional disparity, from 3651 (0.3%) in Qinghai province to 134,318 (11.0%) in Guangdong province. For attitudes, a predominance of positive attitudes was noted across all provinces, ranging from 60.0% to 74.1%, while negative attitudes were less common, ranging from 4.4% to 6.7%. Provinces in the central region exhibited higher prevalence of positive attitudes (72.1% to 74.1%) and lower prevalence of negative attitudes (4.5% to 5.2%), which also showed a relatively higher prevalence of social norms. Conversely, Shanghai, Beijing megacities and three provinces in the northeastern region showed the higher proportion of negative attitudes (5.3% to 6.7%), which also displayed a greater mention of misinformation. Northern regions exhibited higher prevalence than southern regions across all three Health belief constructs (i.e., perceived disease risk, perceived benefits, perceived barriers to accepting vaccines). Central and western provinces, particularly Shaanxi (47.8% of 59,868 posts), reported a significantly higher prevalence of practical barriers associated with HPV vaccine uptake.

**Figure 4.**
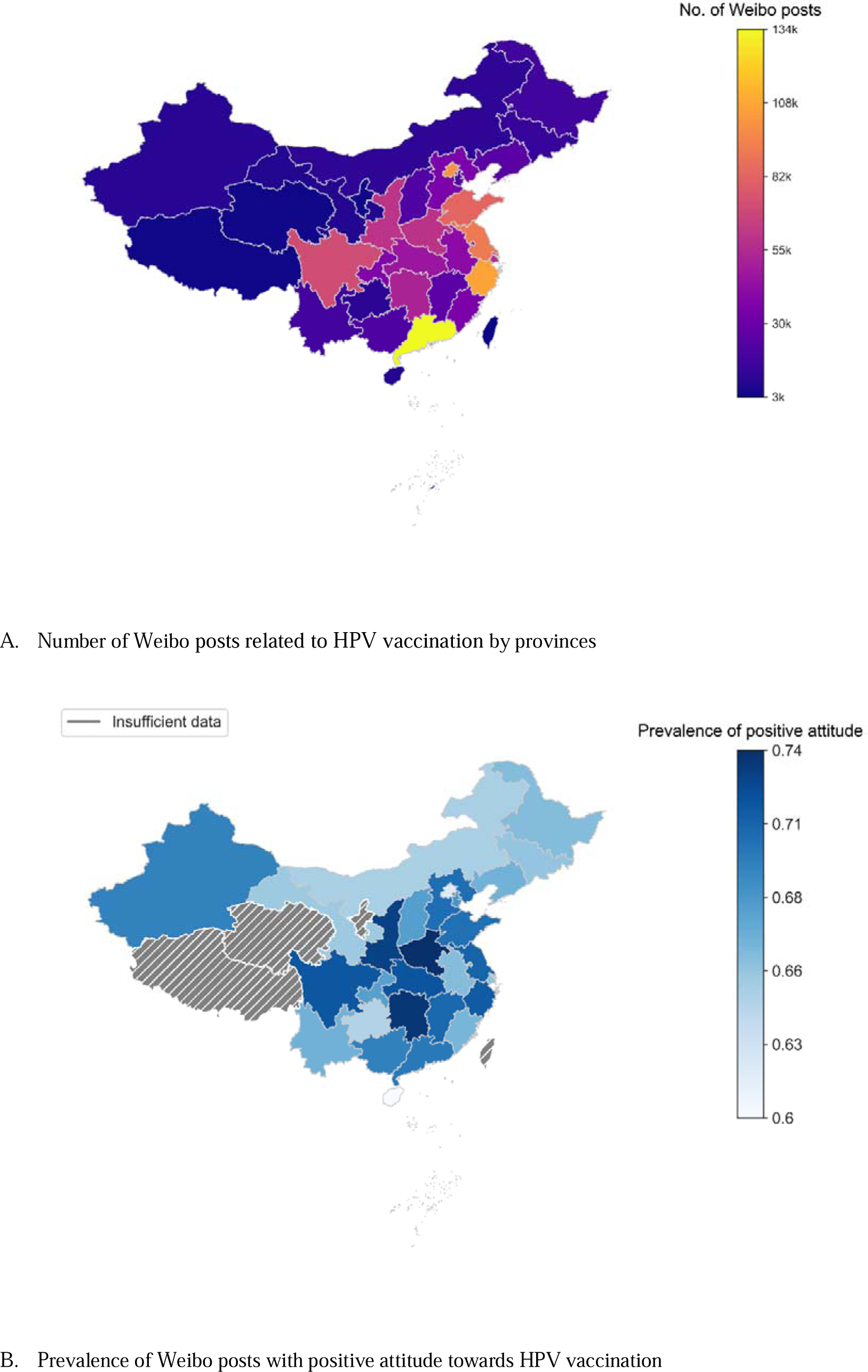
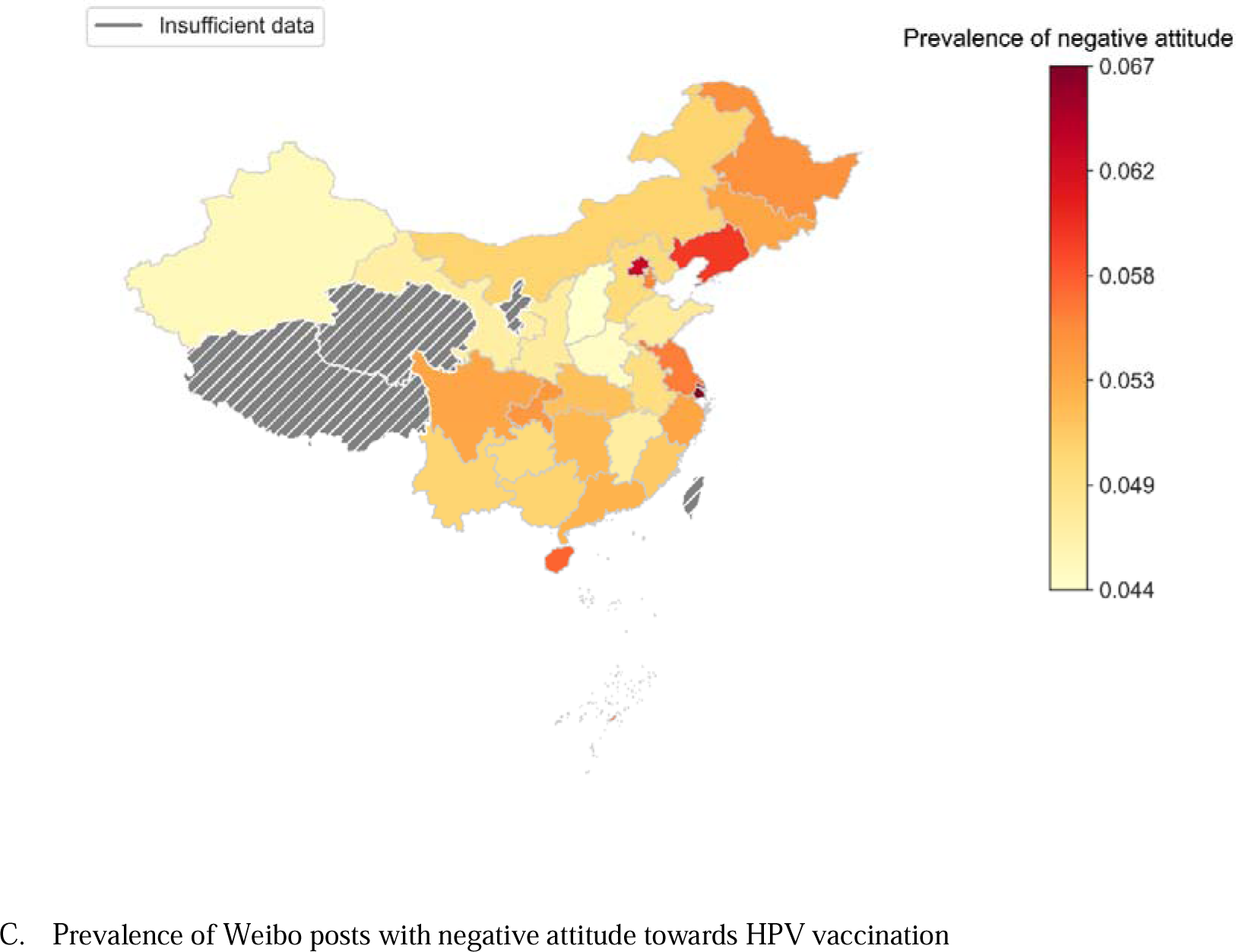
Geographic variations in the number of Weibo posts and the prevalence of posts with positive or negative attitude towards HPV vaccination NOTE: Locally estimated scatterplot smoothing (LOESS) is employed to reveal the temporal trends of prevalence and remove random noise components on monthly raw data.

### Comparison by Types of HPV Vaccines and Gender

Weibo posts related to the 9v vaccine (1,474,618) far outnumbered those to the 2v (26,264) and 4v vaccines (51,538) (eFigure 4, in the supplementary material). The temporal trends by each type of HPV vaccine with smoothing techniques, for the prevalence of attitudes are depicted in Figure 5, and for constructs of health beliefs and environments in the supplementary material (eFigure 5). In total, Weibo users exhibited the most positive attitude to 4v and 9v vaccines (79.6% and 74.1%), which is higher by about 10% than 2v vaccines (65.7%). We also conducted chi-square tests on vaccine type, suggesting significant differences among three types in all framework constructs (p < 0.001). Larger differences by vaccine types were observed for barriers to vaccination instead of facilitators. There are significantly more prevalent on perceived barriers to accepting vaccines (33.5%) and misinformation (16.9%) towards 2v vaccines than 4v (12.1% and 9.4%) and 9v (13.1% and 8.6%) vaccines, but less prevalent on practical barriers to 2v vaccination (eFigure 5, in the supplementary material).

**Figure 5.**
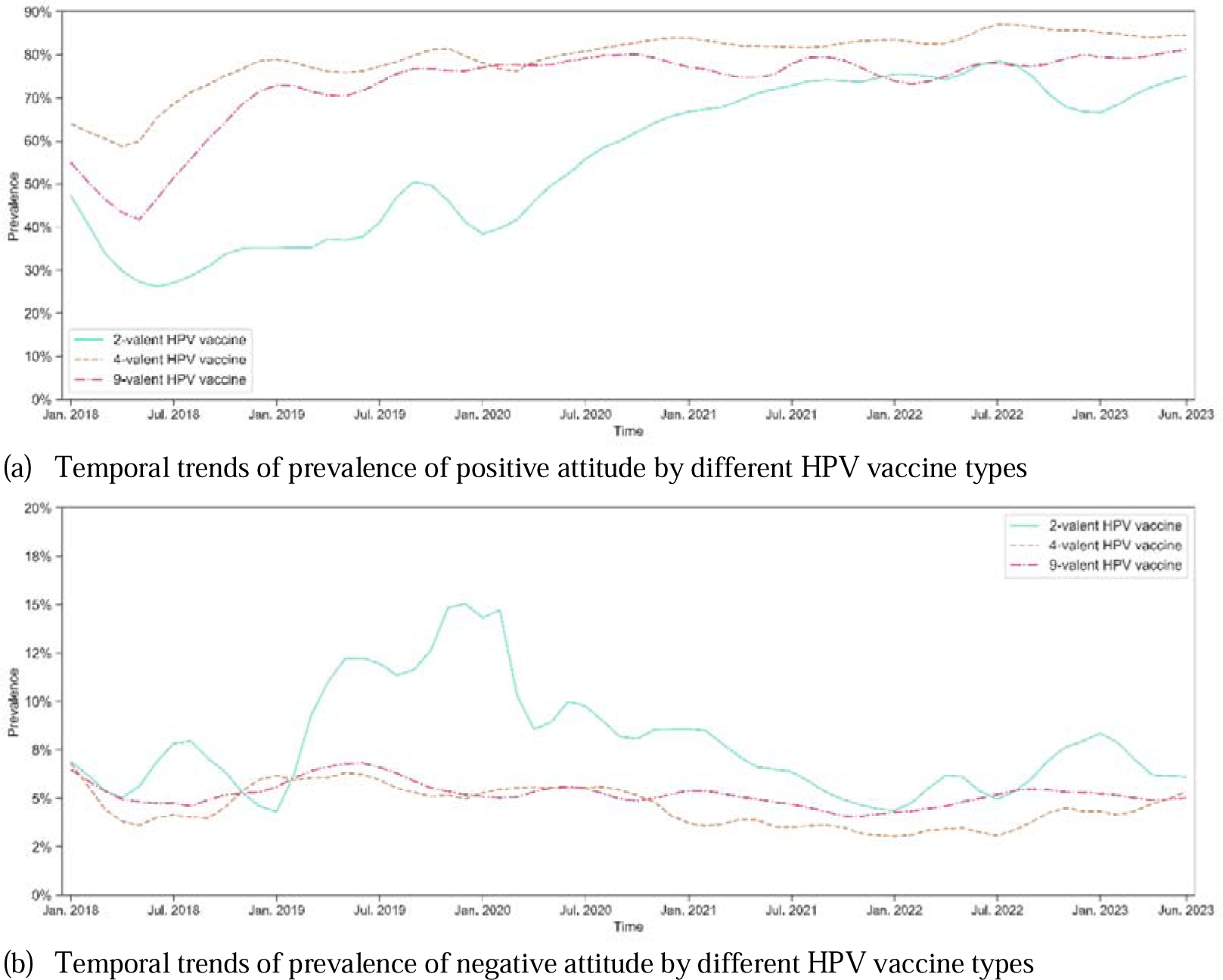
Temporal trends of prevalence of attitudes towards different HPV vaccine types on Weibo

The number of relevant posts from female users (1,533,300) was far greater than those from male users (204,938) (eFigure 6, in the supplementary material). Positive attitude towards HPV vaccination was significantly more prevalent among female than male users (71.5% vs 52.2%).

## Discussion

Social media listening emerged as a promising method to monitor public perceptions on health issues and can enable the development of health communication strategies.^29^ Leveraging deep learning techniques, we analyzed social media posts to capture the temporal and spatial trends in public perceptions, perceived barriers, and facilitators towards HPV vaccination in China according to behavior change theories.^10–12^ Our study would help to develop tailored education strategies and to address the quite low rates of HPV vaccination in China.

Analyzing public attitudes via social media listening can serve as a pivotal tool for identifying prevailing public sentiment, conducting surveillance, and addressing prompt response to anti-vaccine activity. We found that, the prevalence of positive attitudes towards HPV vaccination among Weibo users increased substantially and remained consistently high from 2019 onwards. It contrasted with the situation among social media users in the United States (US), where negative attitudes towards HPV vaccination were more prevalent.^30^

Different to most high-income countries (e.g, US) where only 9v HPV vaccine was supplied, there are three types (2v, 4v, 9v) of HPV vaccines provided in China. However, a sustained higher prevalence of negative attitudes, perceived barriers to accepting vaccines, and misinformation was observed for 2v vaccines in our study. As a domestically developed 2v HPV vaccine demonstrated cost-effectiveness in China, health communication should be promoted to reduce rumors and misinformation, especially for 2v vaccines. Based on gender disparities in the attention and attitudes towards HPV vaccination, we recommend HPV-related health communication targeting males, facilitating a more gender-inclusive environment for vaccine promotions.

On social media, public discussions on HPV disease risk and vaccination benefits displayed a decreasing trend nationwide from early 2018 to mid-2023, which resemble international trends.^22^ After the introduction of HPV vaccination in China, the public attention shifted from discussions on the disease risk and vaccination benefits to practical considerations regarding vaccination decisions. Given the pivotal role of positive health beliefs on disease risks and vaccines, health education should continue to emphasize HPV infection risks and benefits of vaccines before HPV vaccination campaign in city-level or nationwide in China. Furthermore, our study found that practical barriers to vaccination, particularly accessibility issues (e.g., insufficient supply and high price of vaccines), are the most prominent concern among the public, which align with a previous study.^31^ In contrast with high-income countries, improving accessibility is the most paramount to enhancing HPV vaccination coverage in China, and the change in public attitude and social norms should be the first step of policy change. It is imperative for the Chinese government to prioritize addressing the barriers found in social media listening and survey studies.

On Weibo, the prevalence of misinformation and perceived barriers to accepting vaccines decreased throughout the study period, while social norms became more prevalently discussed than misinformation since March 2020. Social norms not only exert a positive influence on intentions of receiving the HPV vaccine,^32^ but also serve as a measure to raise perceived benefits of vaccination and counter misinformation and perceived barriers.^33,34^ Although HPV vaccine-related perceived barriers and misinformation declined in China and were lower than those on platforms like Instagram and X,^35,36^ the importance of managing misinformation should not be ignored. Given the isolated nature of information on social media, misinformation tends to gather and spread rapidly, making it challenging to conquer it.^37^ Health systems can leverage social media listening to identify and track misinformation in real time and facilitate timely intervention, maintaining vigilance to misinformation and mitigate its impact on vaccine perceptions.

In addition, more attention should be paid to regions with a higher prevalence of negative attitudes. Through the geographical analysis, we observed that Beijing, Shanghai, and north-eastern provinces exhibited higher levels of negative attitudes and misinformation. Higher internet penetration rates in megacities like Beijing and Shanghai may contribute to the accelerated spread of misinformation.^38^ Therefore, it is imperative to establish fact-checking mechanisms and promptly prevent the widespread of misinformation. North-eastern regions faced challenges of low health literacy,^39^ which hinders public capacity to obtain health knowledge and combat misinformation.

Compared to conventional survey methods, this study adopted a state-of-the-art DL-based approach to conduct social media listening. This methodology not only enables public health surveillance in real time, but also offers an economically efficient and expeditious means of surveillance, facilitating temporal and geographical analyses on extensive datasets.^16,40^ One advantage of this method is its direct extraction of public discussions from social media platforms, eliminating potential biases stemming from researcher-participant interactions in surveys. Although there is a possible age bias between social media users and the general population, the primary target groups for HPV vaccination are young adults and college students,^41^ which match the age profiles of social media users. These advantages can be leveraged to direct strategic efforts by more effectively informing health communication strategies and tailoring them to specific populations and areas.

Our study also contributes to address a methodological issue reported in previous social media listening research, that performance of DL algorithms would be hurt by training dataset with highly imbalanced label distribution.^42,43^ Since some framework constructs are imbalanced labeled in our social media data, we alleviated this issue to improve the DL performance by integrating an advanced few-shot DL approach (SetFit in our study) and down-sampling technique. The few-shot approach reduces the requirement for manually labeled data and provides the opportunity of designing training dataset using diverse strategies. And down-sampling technique can convert the unbalanced training dataset into a more balanced training dataset. The few-shot approach with down-sampling technique is helpful to process social media data with imbalanced label distribution.

### Limitations

This study has several limitations. First, there may be population bias, as social media users may not represent the broader demographic characteristics of the general population. Social media users tend to be younger, however, exactly aligning with the target population for HPV vaccination.^6^ Second, our analyses are based on individual posts instead of social media users. Future research may develop innovative computational algorithms to analyze at the individual user level by incorporating users’ profiles and historical posts. Third, the gold-standard corpus of 6,600 posts may not fully represent the diversity of the larger unlabeled dataset, which may affect the robustness of the DL prediction. Fourth, although we leveraged an advanced DL approach to mitigate bias possibly induced by label imbalance, there is still a need to improve prediction accuracy.^44^ Nevertheless, DL classifiers in our study demonstrated high accuracy across all tasks, ensuring the reliability in analyses.

### Conclusions

We conducted a deep learning analysis of HPV vaccine-related discussions posted on Weibo platform according to behavior change theories. Our study highlighted the potential of social media listening with DL techniques in public health fields. Time-series analysis revealed the dynamic shifts in public perceptions and exposures to information environments and practical barriers to HPV vaccination. Geographical analyses uncovered regions with a higher prevalence of negative attitudes and misinformation. Our findings would help to develop tailored education strategies and to improve HPV vaccination in China.

### Ethical statement

The study was approved by the Institutional Review Board of the School of Public Health, Fudan University (IRB#2022-01-0938).

## Contributors

ZH conceived the study and led the analysis. YW, ZD, YZ and LM completed the manual annotation. HY implemented data collection, data pre-processing, deep learning analysis, and data visualisation. YW implemented statistical analyses. XZ and ZH verified the data. YW, ZD and ZH wrote the manuscript. All authors contributed to reviewing and editing of the manuscript. ZH had full access to all the data in the study and made the final decision to submit for publication.

## Data sharing statement

All data described in the results and corresponding Python/R codes will be shared on <u>GitHub</u> upon acceptance of this paper. Original posts are not shared according to Weibo’s data policy. Other data are available on request from the corresponding author.

## Declaration of interests

ZH received funding from Merck Investigator Initiated Studies. The other authors declare no competing interests.

## Data Availability

All data described in the results and corresponding Python/R codes will be shared on GitHub upon acceptance of this paper. Original posts are not shared according to Weibo's data policy. Other data are available on request from the corresponding author.

## Acknowledgements

ZH acknowledges financial support from Merck Investigator Initiated Studies (61185). The funders had no role in the study design, data collection, data analysis, data interpretation, or writing of the report.

## Notes

### Author Declarations

The study used ONLY openly available human data that were originally presented at Weibo social media platform. The study was approved by the Institutional Review Board of the School of Public Health, Fudan University (IRB#2022-01-0938).

