## Supplementary Materials for "Trajectories of and spatial variations in HPV vaccine discussions on Weibo, 2018-2023: a deep learning analysis"

### Supplemental Methods

**Development of Annotation Framework**

We developed our annotation framework and defined constructs in the framework on the basis of Health Belief Model (HBM), Theory of Planned Behavior (TPB), and Increasing Vaccination Model. Grounded in the HBM, our framework identified key factors of HPV vaccination, including perceived susceptibility, severity, benefits, and barriers, which shape individuals’ internal health beliefs and drive health-related attitudes, serving as primary determinants of behaviors.^1,2^ Due to a limited corpus of relevant social media posts, we combined perceived susceptibility and severity into the broader construct “perceived disease risk”. Our framework also considered environmental information processes that could influence attitudes toward HPV vaccination, focusing on constructs of positive social norms and misinformation, which align with the nature of vaccination-related behavior and embody the characteristics of social media content.^3,4^ Within the TPB framework, our framework centered on the construct of attitudes, comprising fact-based statements, inclusive of reference to norms and/or behavioral control but defined by positive, negative, and neutral valence.^5^ Additionally, we examined practical barriers to vaccination discouraging individuals with positive intentions from actual uptake.^6,7^ Although self-efficacy/perceived behavioral control, an individual’s perception of his or her competence to successfully address practical issues and perform a behavior, is another crucial concept in both HBM and TPB frameworks, we did not set a corresponding construct to capture it in our framework deliberately, since it can hardly be accurately reflected and measured in terms of social media languages. Nevertheless, since another construct of our framework, practical barriers to vaccination, measures the user-reported exposures to practical issues preventing people willing to get vaccinated from conducting vaccination behaviors, it can be regarded partially as an opposite measure of perceived control,^6^ ensuring integrity of our conceptual framework.

In addition, we eliminated the posts from official accounts, propaganda or news reports, assigning them to category “irrelevant” with those irrelevant to HPV vaccination, in order to present the perceptions of Weibo users. We sought to enhance clarity by providing comprehensive definitions of the included constructs and illustrative raw examples of corresponding posts from the social media platform, which are presented in eTable 1.

**eTable 1.** Definitions and Examples of Constructs in the Annotation Framework

| **Construct and category** | **Definition** | **Sample raw posts (translated into English by authors)** |
| --- | --- | --- |
| Irrelevant | Posts by official accounts, media accounts, news reports, and propaganda or non-HPV vaccine related posts, or forwards (//) without indicating the opinions or sentiments of the forwarder. | - //@BigTycoon: Do all girls really need the nine-valent one? - #XiaomiMIXFold2Valent# Been keeping an eye on this for a while, and it's finally here. The ultra-slim design, paired with its hinge mechanism, makes it truly stand out from the crowd. - #NineValentHPV #NineValentVaccine Book your nine-valent vaccine appointment in Zhoukou, Zhumadian, Wuhan, Huangshi - appointments available, with scheduling taking approximately one week to a month. |
| Positive attitude | Positive opinion or prompt HPV vaccine. | - By the way, before I dive into a relationship and say ‘I do’ 👰🏻, I’m making sure to get my full nine-valent vaccine, better safe than sorry! |
| Neutral attitude | Related to HPV vaccine topic but contains no sentiment or sentiment is unclear. | - HPV vaccine, mobile phone |
| Negative attitude | Concerns, doubts, hesitancy, or refusal about the HPV vaccine, perceptions of intense side effects or safety issues. | - Give me another chance, and I promise I won't opt for the nine-valent HPV vaccine. Regret, regret, regret, regret, regret, regret, regret, regret, regret, regret, regret. |
| Perceived disease risk | Subjective evaluation of the risk or severity of getting an HPV infection, related disease, or its potential consequences. | - #NineValentHPVVaccine# Gotta save up some cash 💰 for those three shots of nine-valent! [LOL] Don't want to get sick! |
| Perceived benefits of vaccines | Individual’s confidence in the value, efficacy, or necessity of the HPV vaccine in protecting against HPV infection, HPV infection-induced cancers, and so forth. | - If you have the means, definitely go get the HPV vaccine, it really works!! |
| Perceived barriers to accepting vaccines | Lack of confidence in vaccination, feeling skeptical about the safety and efficacy of the vaccine, worried about side effects and discomfort (e.g., pain, emotional upset), inaccurately concerned about the possible harms or risks associated with vaccination. | - After getting the first dose of the 9-valent vaccine, I did a quick side effects search, and it really made me reconsider the next two shots. |
| Social norms | Exposure to external facilitators, such as events or information from close others, the media, an important person or group, or health care providers promoting HPV vaccination, that trigger or motivate individuals to accept HPV vaccination. | - HPV vaccinations seem to be popping up everywhere! Is this a sign for me? |
| Misinformation | Exposure to false, inaccurate or negative information on HPV infection and vaccines, such as rumors, anti-vaccine, vaccine inefficacy, vaccination postponement, alternative medicine, civil liberties, conspiracy theories, falsehoods, negative news and reports, and negative recommendation. | - When it comes to HPV nine-valent vaccine side effects, it feels like only a handful know the real deal, while the rest of us are just brainwashed by the capitalist media wave, trying to stay trend |
| Practical barriers to vaccination | Perceived practical, health services-based issues related to the delivery of vaccination services for individuals intending to uptake to actual accessing vaccines, including lack of vaccine supply, prolonged queue time, inconvenience, expense, and low quality of services. | - Getting that nine-valent vaccine is such a challenge, and the hurdles don’t end even after finally securing an appointment. It’s beyond frustrating! |

**Manual Annotation**

Manually annotated gold-standard corpus is essential to fine-tune deep learning (DL) algorithms for analyzing HPV vaccine-related posts. We performed random extraction of 100 posts per month within the observation period, resulting in a subset of 6,600 posts in total for manual annotation.

The manual coding procedure for social media posts was executed in accordance with established research methodologies to ensure methodological rigor.^5^ Two annotators independently annotated each of 6,600 posts according to its relevance to the constructs in the annotation framework. Each post was categorized as either unrelated to any of the constructs or associated with one construct or more. For the category of attitudes, the annotators identified the overall attitude expressed by each post toward the HPV vaccine and assigned them as positive, negative, or neutral. If a post was categorized as “Irrelevant” during annotation, it was excluded from further consideration against other constructs. The initial 400 posts were used for annotation training purposes, with the annotators independently annotating each post and subsequently engaging in group discussions to reach a consensus or majority vote decision on all posts. Training concluded after group consensus was reached on all posts. Following the annotation training, the remaining sampled Weibo posts (6,200) were distributed among two annotators for independent coding. To assess the reliability of the annotation process, we then computed Cohen’s κ among two reviewers.^8^ After the calculation, consensus was also achieved to resolve any coding discrepancies that arose during the independent coding. Ultimately, this meticulously curated gold-standard corpus of totally 6,600 manually annotated posts served as the foundation for fine-tuning and evaluating the DL algorithms. Cohen’s κ interrater agreement was 0.933 for attitudes categorization, and ranged from 0.846 to 0.980 for each non-attitude construct (see eTable 2).

**eTable 2.** Inter-rater agreement of each construct in the Annotation Framework

| **Framework construct** | **Cohen’s κ** | **Standard Error** | **95% CI** |
| --- | --- | --- | --- |
| Irrelevant | 0.97872 | 0.00613 | 0.96671 to 0.99074 |
| Attitudes | 0.93276 | 0.01123 | 0.91076 to 0.95476 |
| Perceived disease risk | 0.87947 | 0.02544 | 0.82961 to 0.92933 |
| Perceived benefits of vaccines | 0.88674 | 0.01749 | 0.85247 to 0.92101 |
| Perceived barriers to accepting vaccines | 0.84581 | 0.02187 | 0.80294 to 0.88868 |
| Social norms | 0.93331 | 0.00934 | 0.91499 to 0.95162 |
| Misinformation | 0.89479 | 0.01842 | 0.85868 to 0.93089 |
| Practical barriers to vaccination | 0.93518 | 0.00673 | 0.92198 to 0.94837 |

**Fine-tuning Deep Learning Models for Classification**

Applying DL models to process text classification, high imbalanced distribution of classification labels in training dataset will hurt the predictive performance of DL algorithms.^9^ Since the relevance of social media posts with most of our theoretical constructs (eg., misinformation, perceived disease risk) was small (about 10%), we needed to resolve the issue of using highly imbalanced posts labeled as a training set. One common and effective way is to leverage down-sampling, which in our case, is to deliberately reduce the volume of majority labels for each training cycle, limiting the ratio between the numbers of input posts labeled differently and not exceeding 2:1. Therefore, to maintain the input data for each training cycle at an appropriate level, we chose to use a few-shot approach requiring only a relatively small sample of data for training process – among which the SetFit (Sentence Transformer Fine-tuning) model is the most advanced one to date. ^10,11^

The SetFit model is based on the Sentence Transformer (ST) model.^11^ The ST model requires the use of a pre-trained BERT (Bidirectional Embedding Representation Transformer) model for encoding, which in our study, was the DMetaSoul/sbert-chinese-general-v2 model.^12^ It is more balanced, has more prominent generalization ability in multiple tasks, and can be directly transformed into a ST model. We developed specific classification model for each framework construct. In the training process, we used Optuna library which is built-in in ST to conduct 10 optimal hyperparameter tunings, and the search spaces are as following:

a. “learning_rate”: set as a floating-point number with log scale, value on the interval [1e-6, 1e-4],
b. “num_epochs”: 1 or 2,
c. “batch_size”: 16 or 32,
d. “seed”: an integer on the interval [1, 40],
e. “num_iterations”: a parameter selected from 5, 10 and 20
f. “max_iter”: an integer on the interval [50, 300],
g. “solver”: one selected from the three methods [“newton-cg”, “lbfgs”, “liblinear”]

We adopted a two-stage hierarchical classification strategy to fine-tune multiple DL classification models (classifiers) using 6,600 manually annotated posts as gold-standard corpus. The first stage is a fine-tuned DL classifier to distinguish whether a post is related to the HPV vaccine or not, and the second stage includes seven DL classifiers for labeling HPV vaccine-relevant posts to each of all seven constructs in the annotation framework. Of the 6,600 sampled posts, we first manually labeled 1,000 posts as relevant to HPV vaccines or not, with which we trained the first stage DL classifier; the first stage DL classifier subsequently classified the remaining 5,600 posts as relevant to HPV vaccines or not. Posts classified as HPV vaccine-related in the first stage were then used for manual annotation against each framework construct and to train the second stage DL classifiers using down-sampling techniques. For each DL classifiers, we randomly selected 80% of the manually annotated data as the training set and reserved 20% as the test set, and all the DL classifiers achieved an accuracy over 0.78 (Table 1).

For the evaluation of SetFit models, we used the methods from the Sklearn library.^13^ We set average = “binary” for the six constructs of health beliefs and environments containing two values. Only one exception is the framework construct of attitudes which have three values (positive, neutral, and negative), the binary strategy cannot be used. We then provide an evaluation of the classification performance on the attitudes construct in detail, see eTable3. The 95% confidence intervals of the classification results are calculated using the bootstrap method.

**eTable 3.** Predictive performance of SetFit model on attitudes towards HPV vaccination in Weibo posts

| **Attitudes** | **Predictive performance of SetFit model** | | | |
| --- | --- | --- | --- | --- |
|  | **Support** | **Precision** | **Recall** | **F-1 score** |
| Positive | 59 | 0.88 | 0.88 | 0.88 |
| Negative | 26 | 0.68 | 0.50 | 0.58 |
| Neutral | 44 | 0.69 | 0.80 | 0.74 |
| Accuracy | 129 |  |  | 0.78 |
| Macro average | 129 | 0.75 | 0.73 | 0.73 |
| Weighted average | 129 | 0.78 | 0.78 | 0.77 |

### Supplemental Results

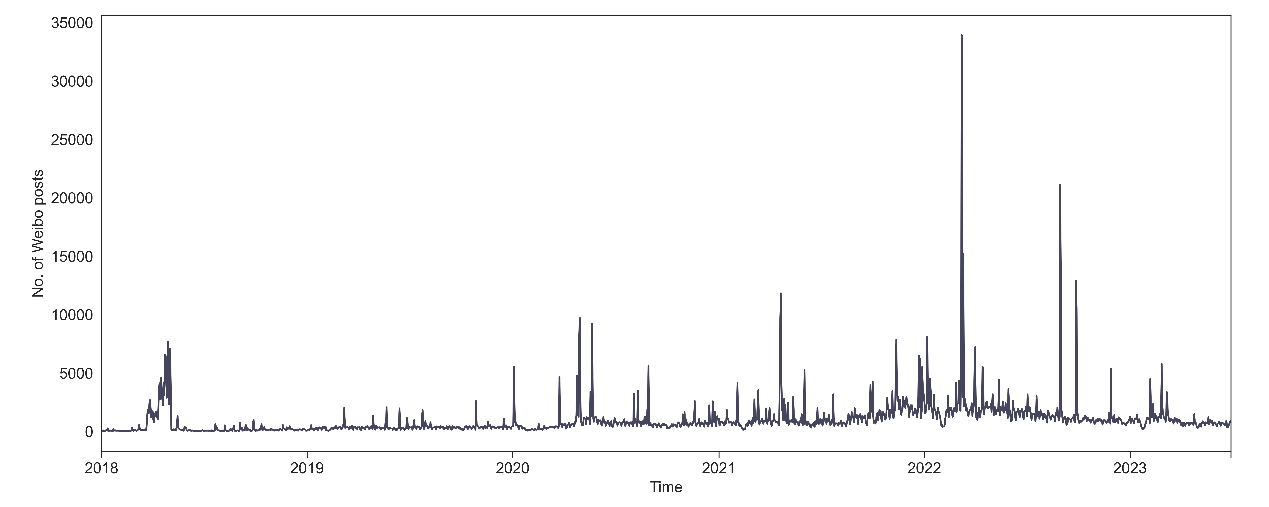

(a) Daily number of Weibo posts related to HPV vaccination

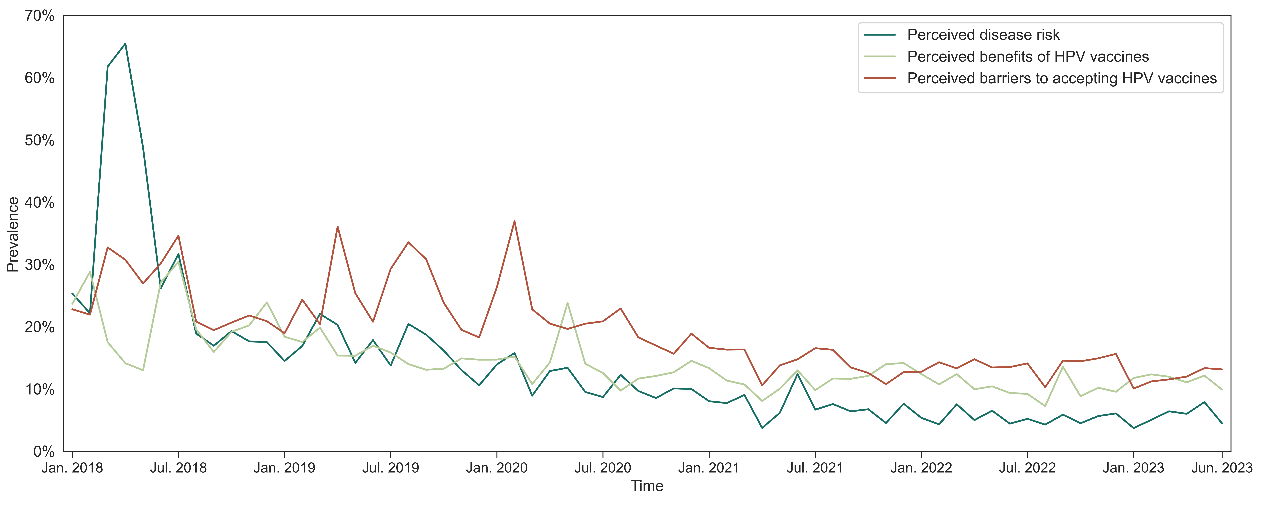

(b) Raw data of monthly prevalence of Weibo posts related to each health belief construct

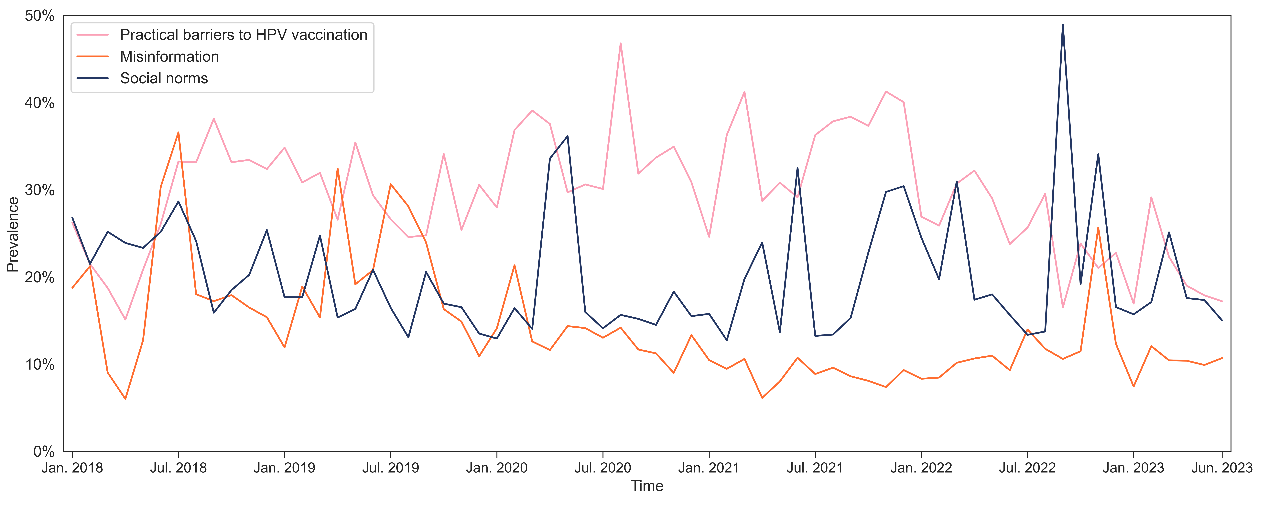

(c) Raw data of monthly prevalence of Weibo posts related to each environment construct

NOTE: Health beliefs indicate constructs of individual’s internal beliefs about HPV and HPV vaccine, including perceived disease risk of HPV, perceived benefits of vaccines and perceived barriers to accepting vaccines; environments indicate constructs of individual’s exposure to environmental barriers or facilitators regarding HPV vaccination, including social norms facilitating HPV vaccination, misinformation about HPV or HPV vaccines, and practical barriers to vaccination.

Different from Figure 3 in Results, these figures show the raw data without smoothing techniques identifying temporal trends.

**eFigure 1.** Raw data of the daily number of Weibo posts and monthly prevalence of posts related to each framework construct of HPV vaccination

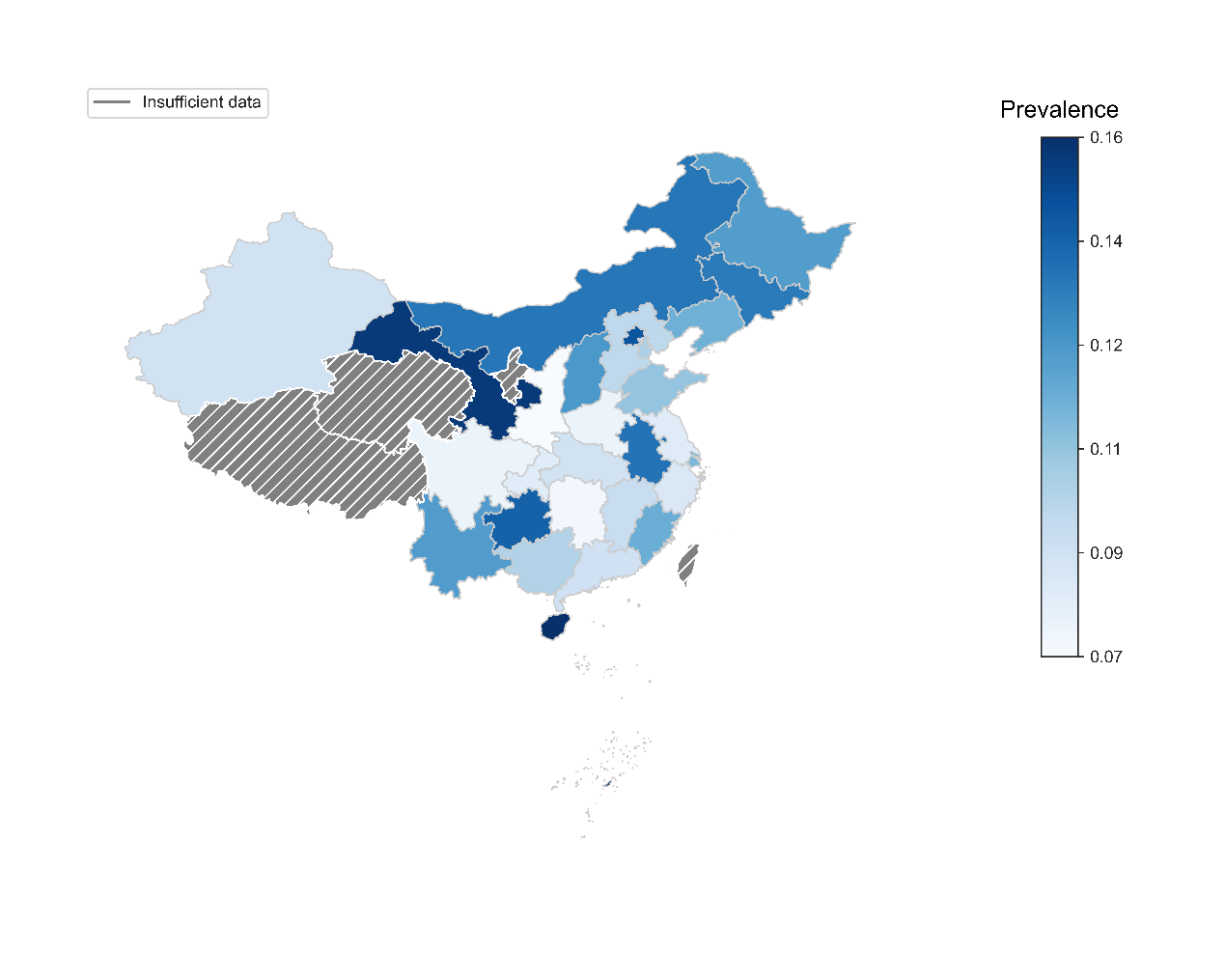

(a) Prevalence of Weibo posts related to perceived disease risk of HPV

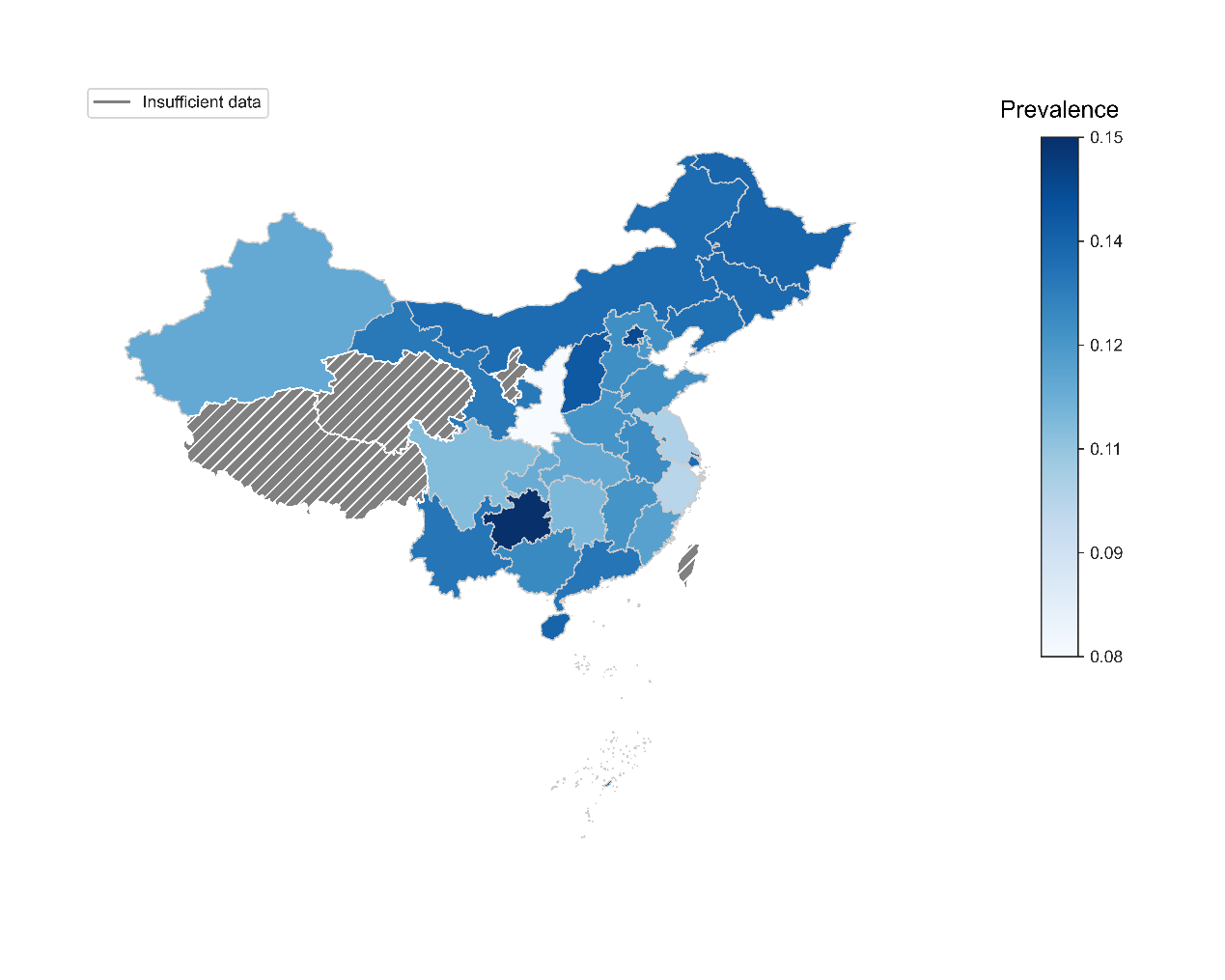

(b) Prevalence of Weibo posts related to perceived benefits of HPV vaccines

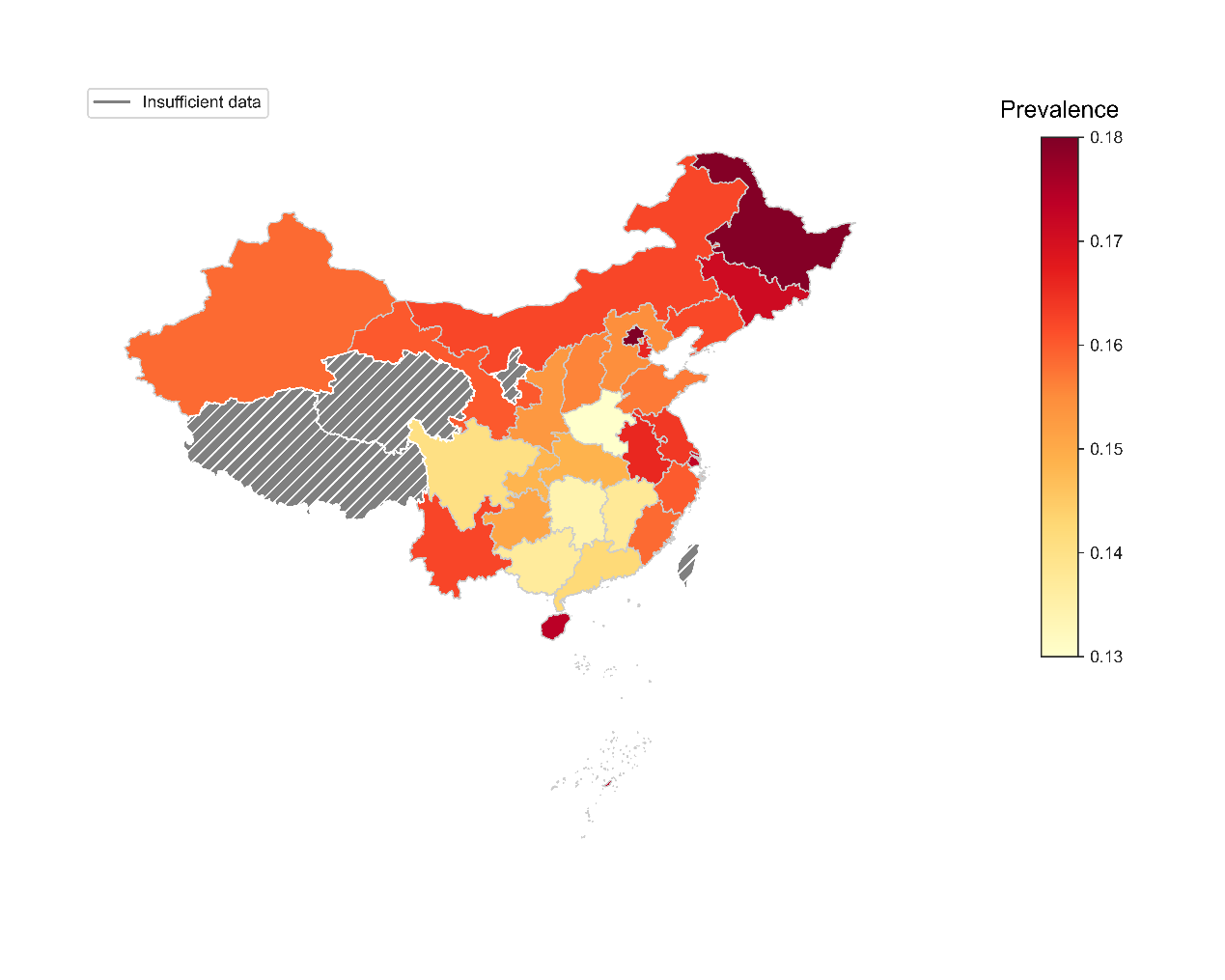

(c) Prevalence of Weibo posts related to perceived barriers to accepting HPV vaccines

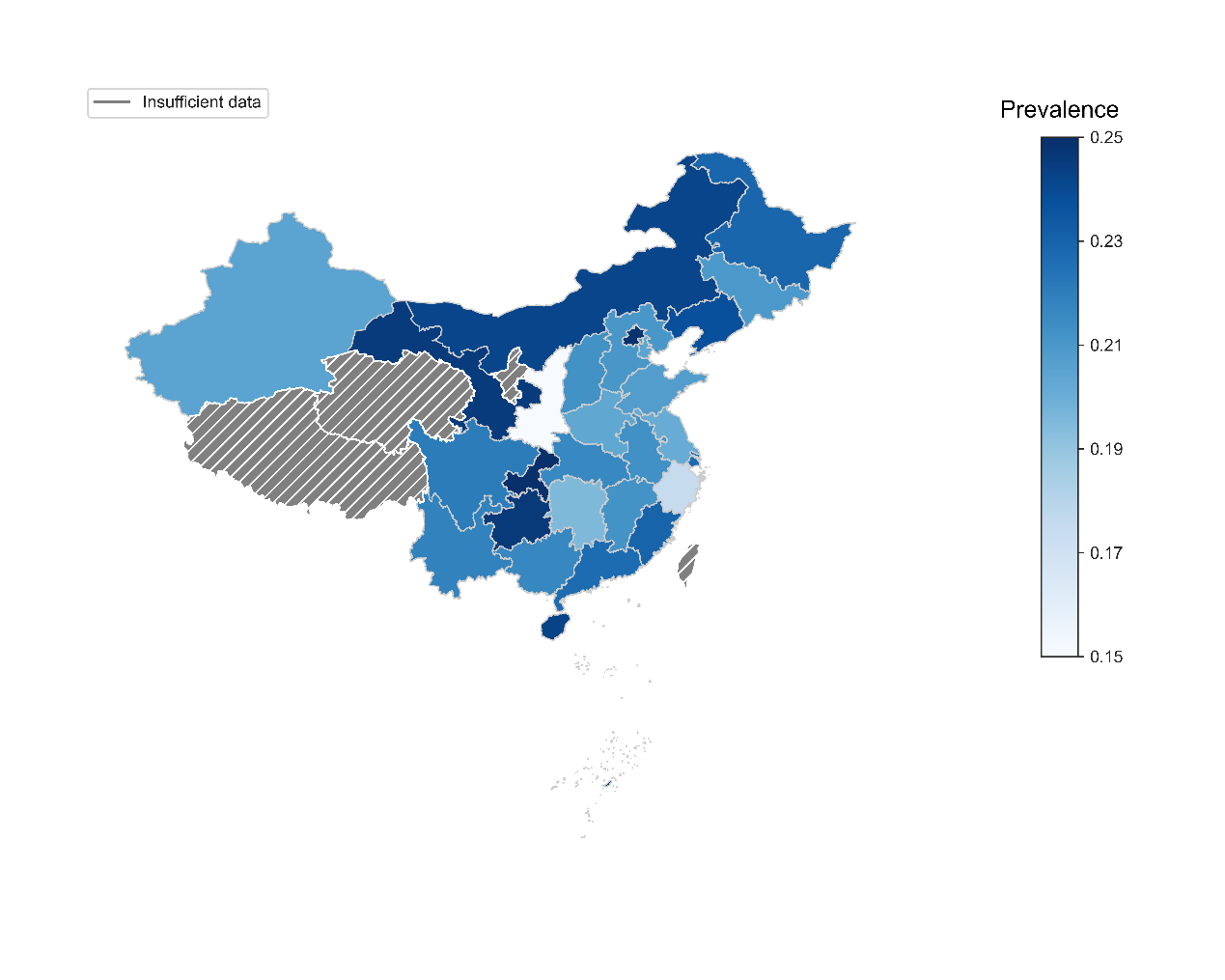

(d) Prevalence of Weibo posts related to social norms facilitating HPV vaccination

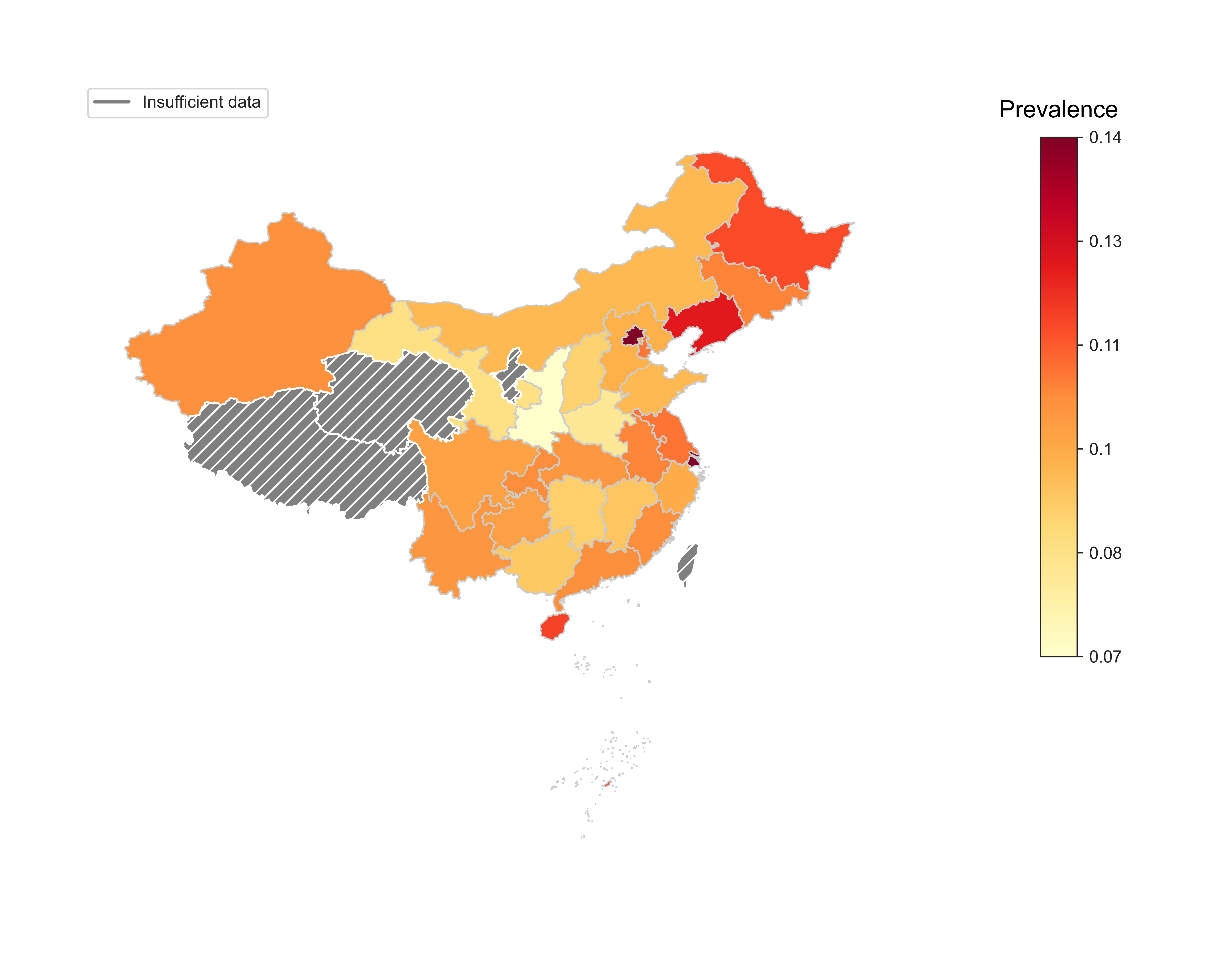

(e) Prevalence of Weibo posts related to misinformation about HPV and HPV vaccines

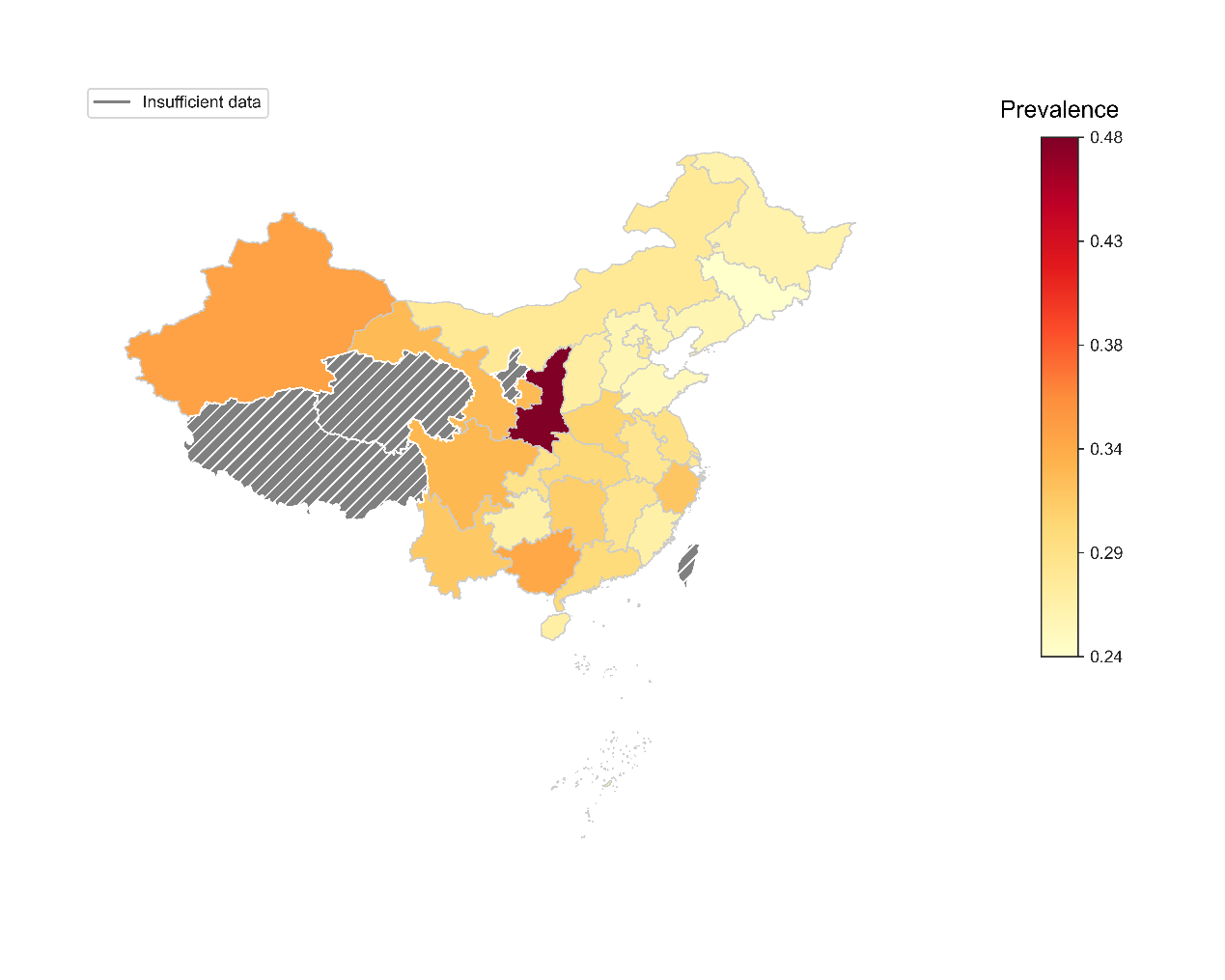

(f) Prevalence of Weibo posts related to practical barriers to HPV vaccination

NOTE: Insufficient data indicates regions excluded from geographic analyses: 1) Hong Kong, Macau, and Taiwan because of different HPV vaccination policy from mainland China; 2) Qinghai, Tibet, and Ningxia (3651, 3709, and 3749 posts) because of too small sample size.

**eFigure 2.** Geographic variations on the prevalence of Weibo posts related to health beliefs and environments of HPV vaccination

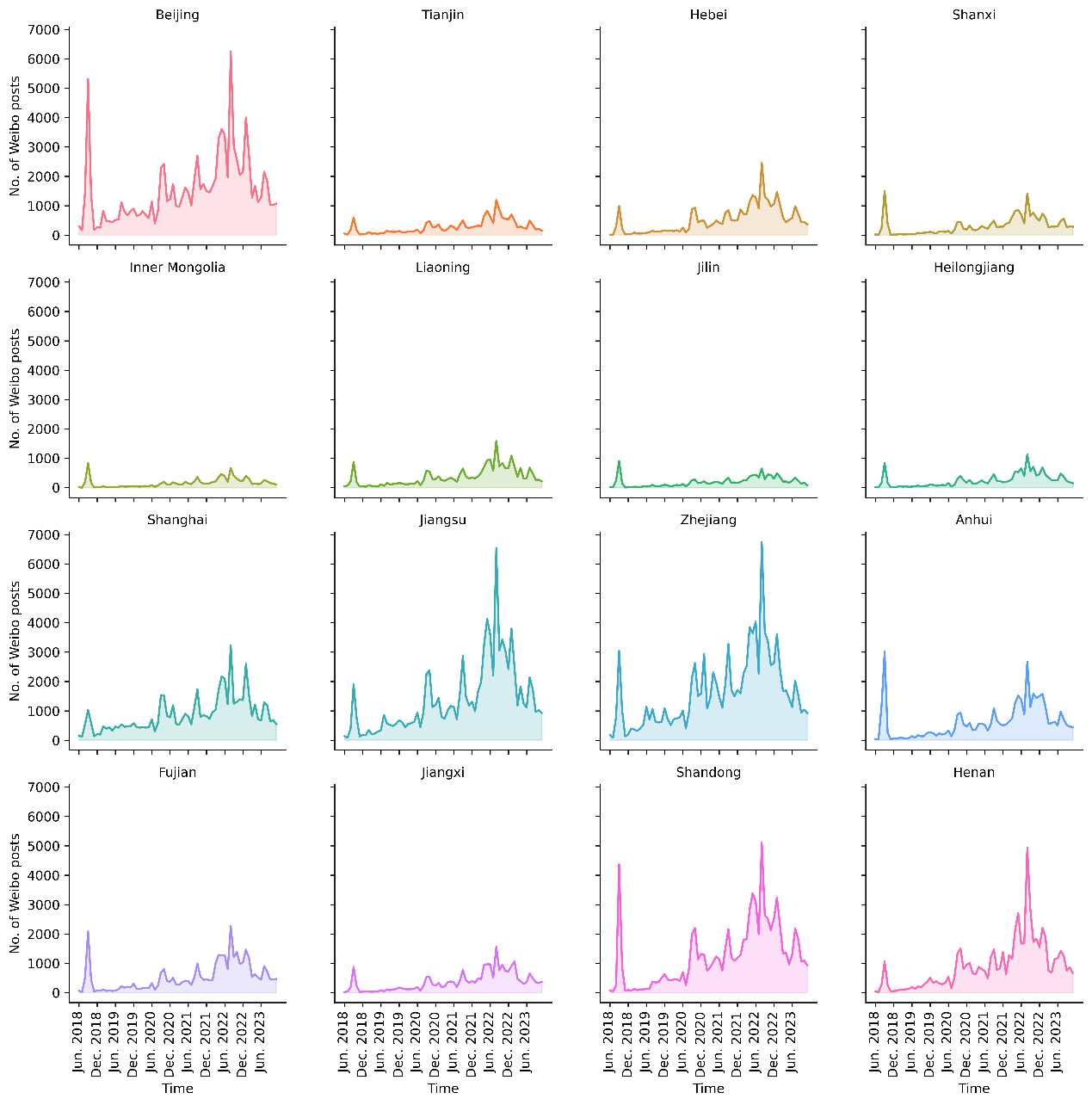

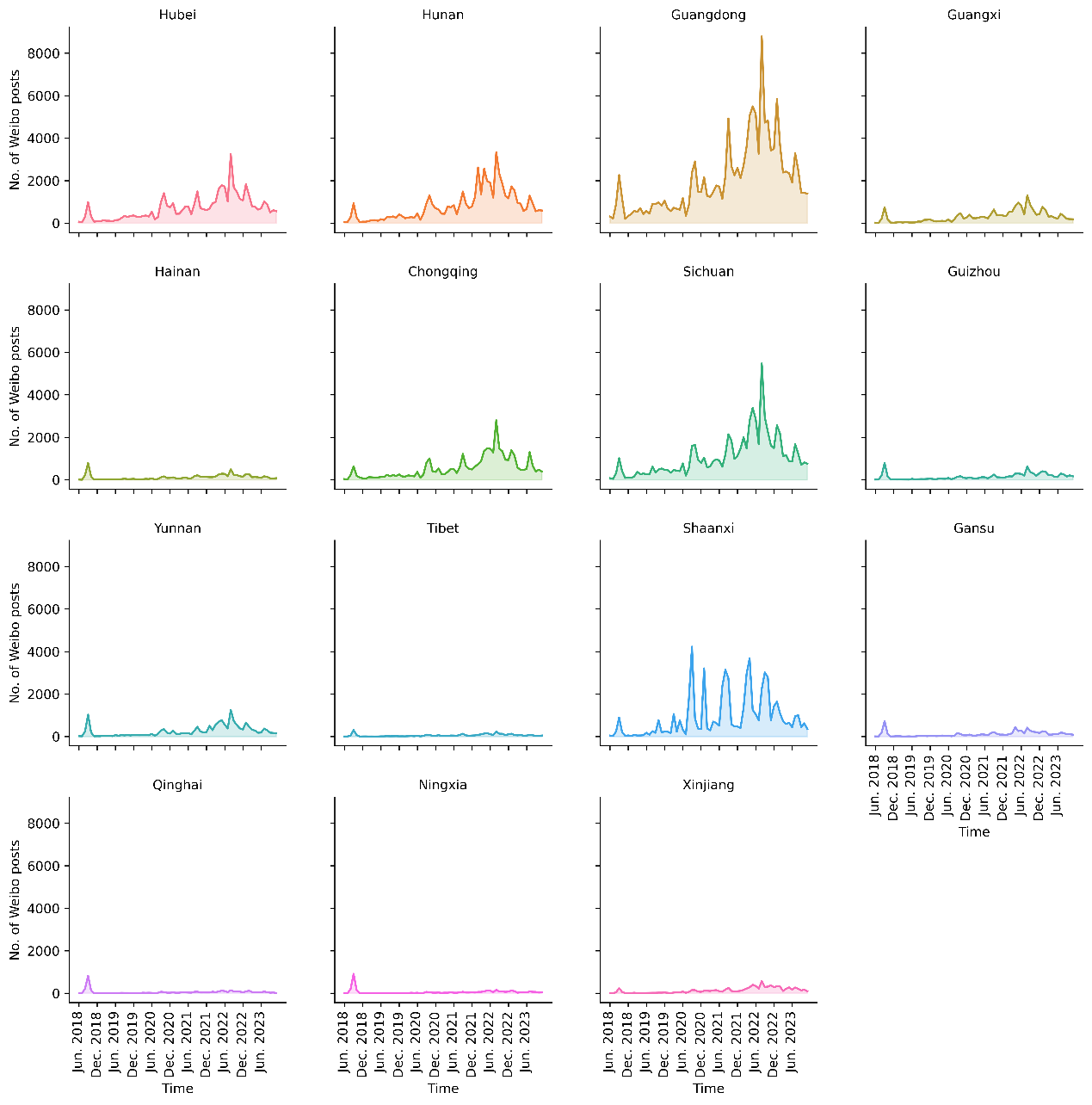

**eFigure 3.** Monthly number of Weibo posts related to HPV vaccination by province

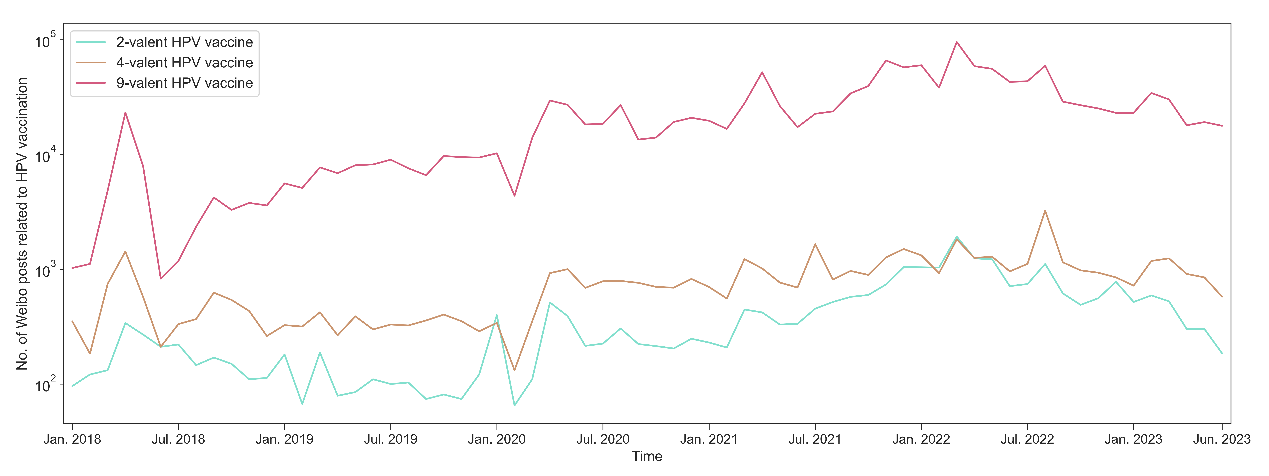

**eFigure 4.** Monthly number of Weibo posts related to HPV vaccination by relevance to different HPV vaccine types

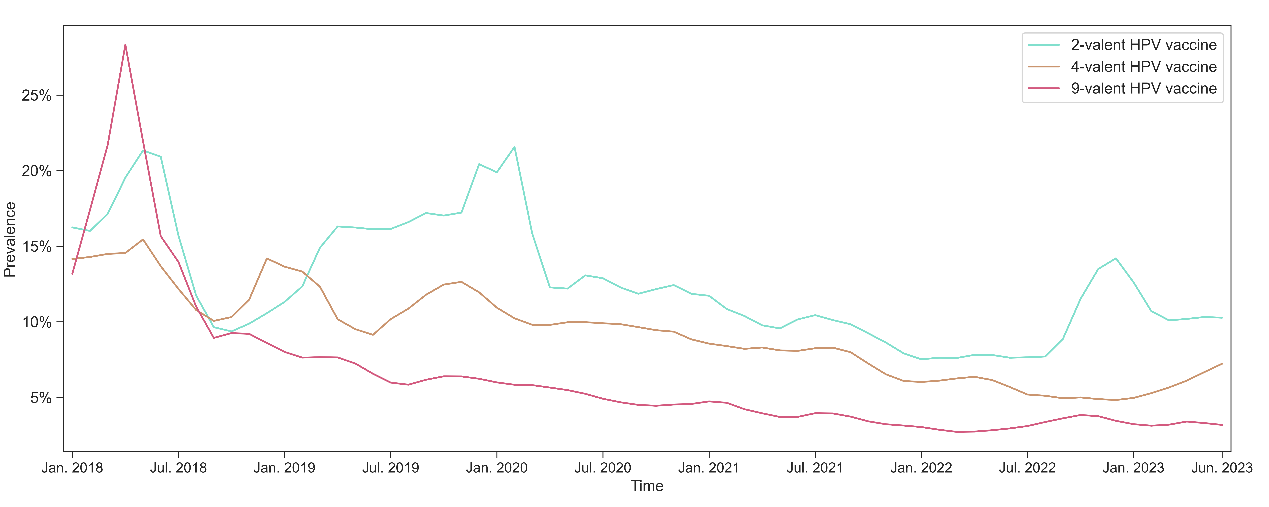

(a) Temporal trends of prevalence of Weibo posts related to perceived disease risk by HPV vaccine types

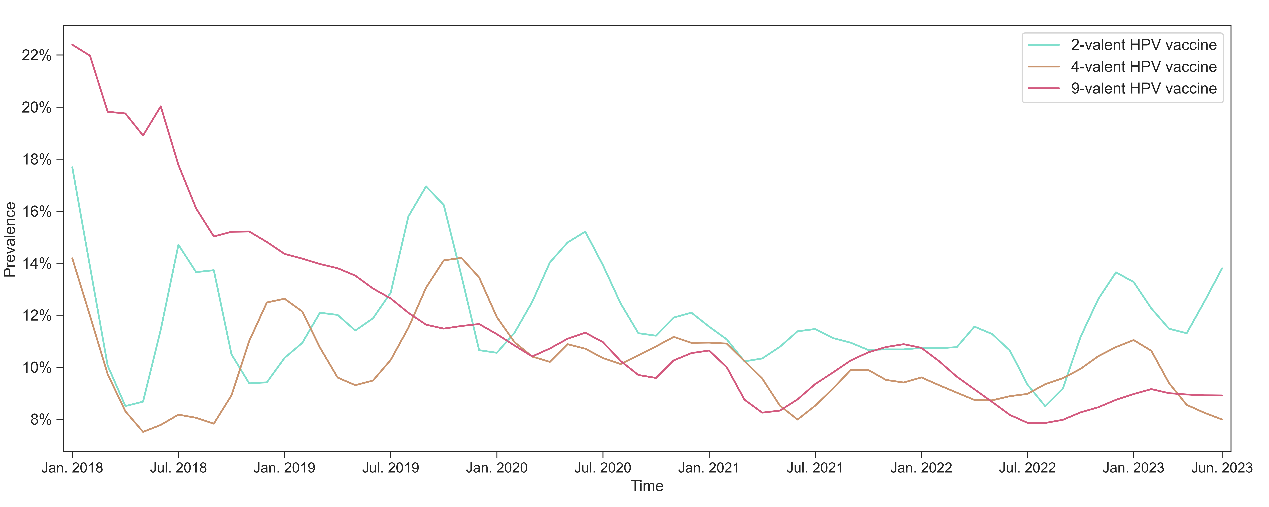

(b) Temporal trends of prevalence of Weibo posts related to perceived benefits by HPV vaccine types

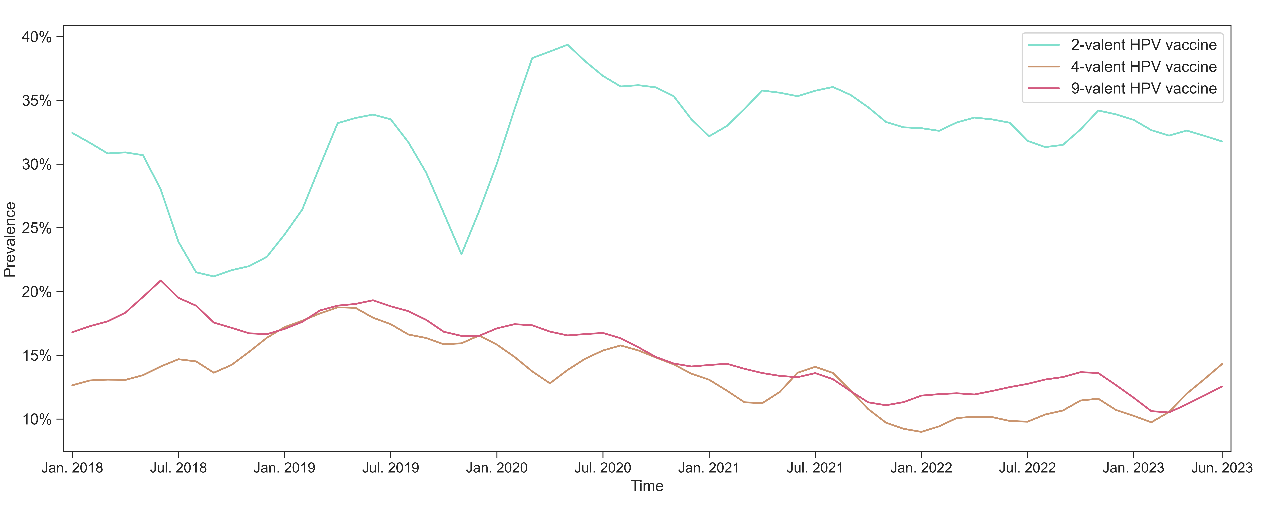

(c) Temporal trends of prevalence of Weibo posts related to perceived barriers to accepting vaccines by HPV vaccine types

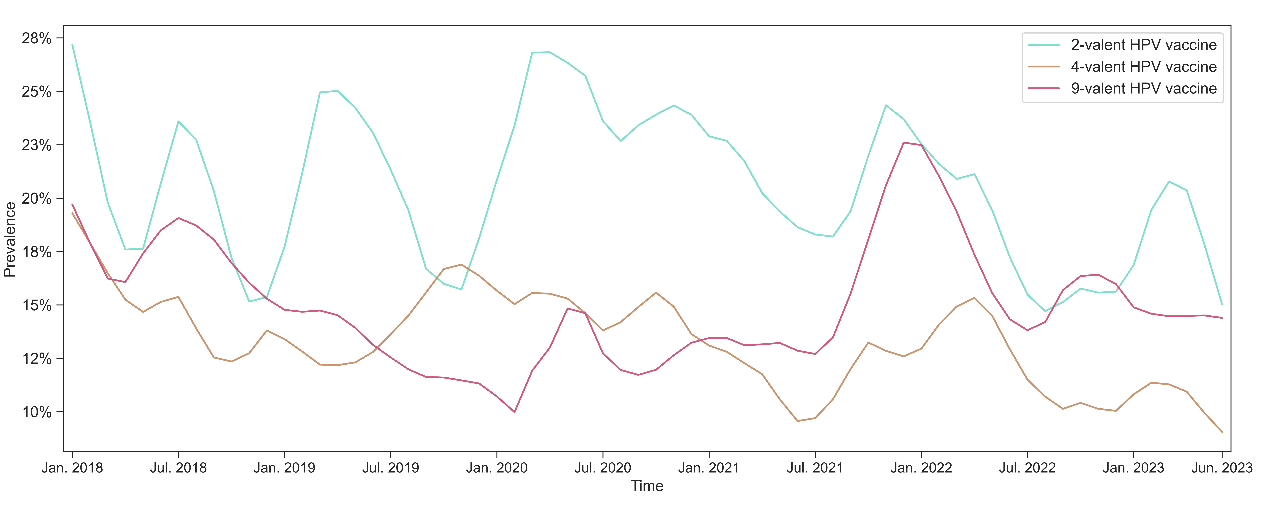

(d) Temporal trends of prevalence of Weibo posts related to social norms by HPV vaccine types

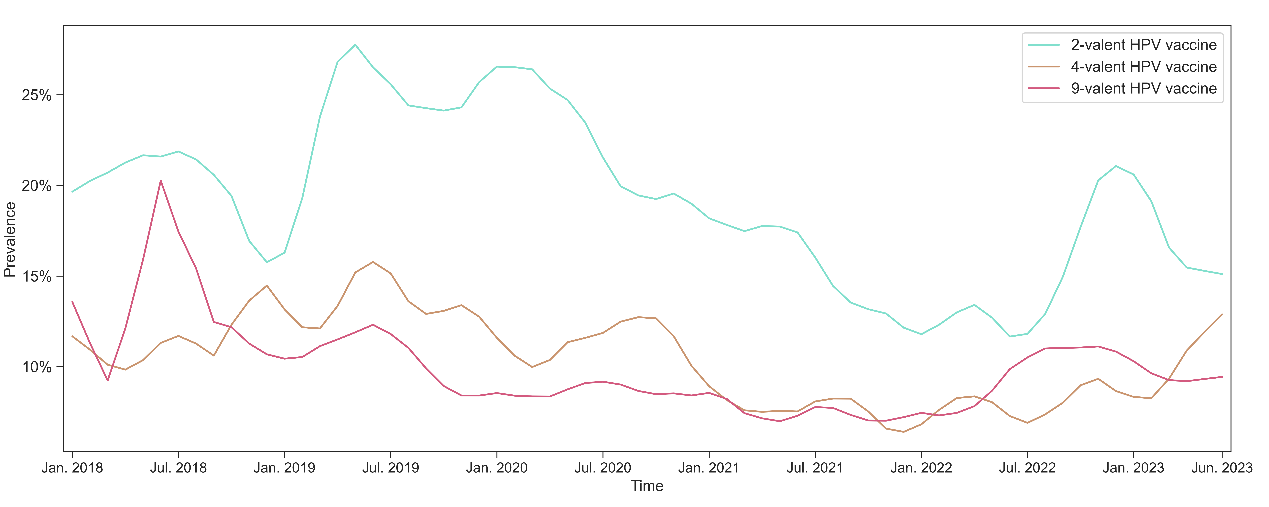

(e) Temporal trends of prevalence of Weibo posts related to misinformation by HPV vaccine types

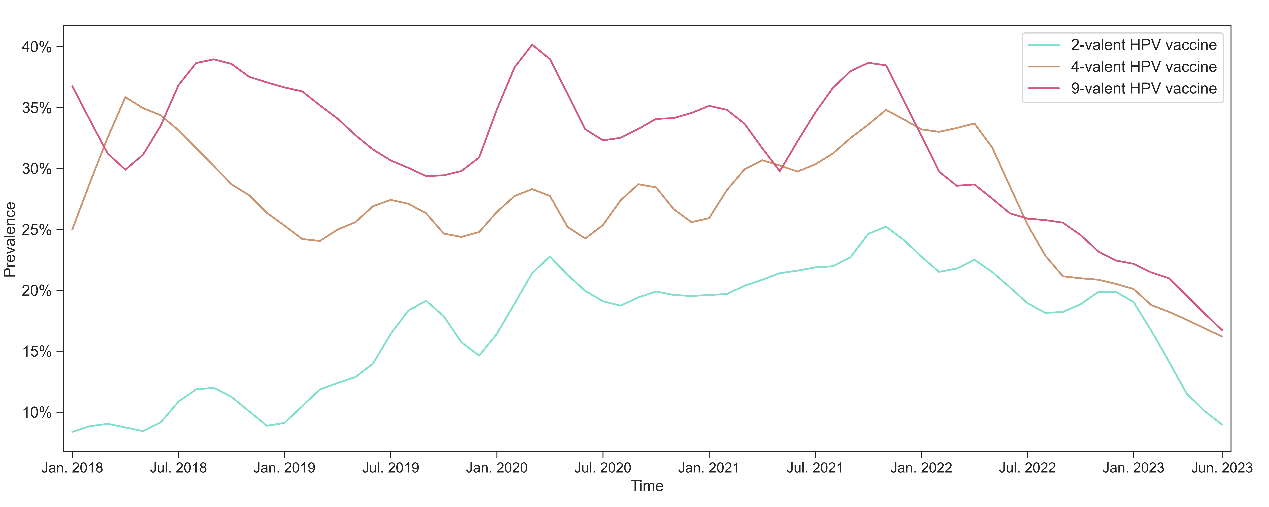

(f) Temporal trends of prevalence of Weibo posts related to practical barriers to vaccination by HPV vaccine types

NOTE: Locally estimated scatterplot smoothing (LOESS) is employed to reveal the temporal trends of prevalence and remove random noise components on monthly raw data.

**eFigure 5.**  Temporal trends of the prevalence of Weibo posts related to health beliefs and environments by different HPV vaccine types

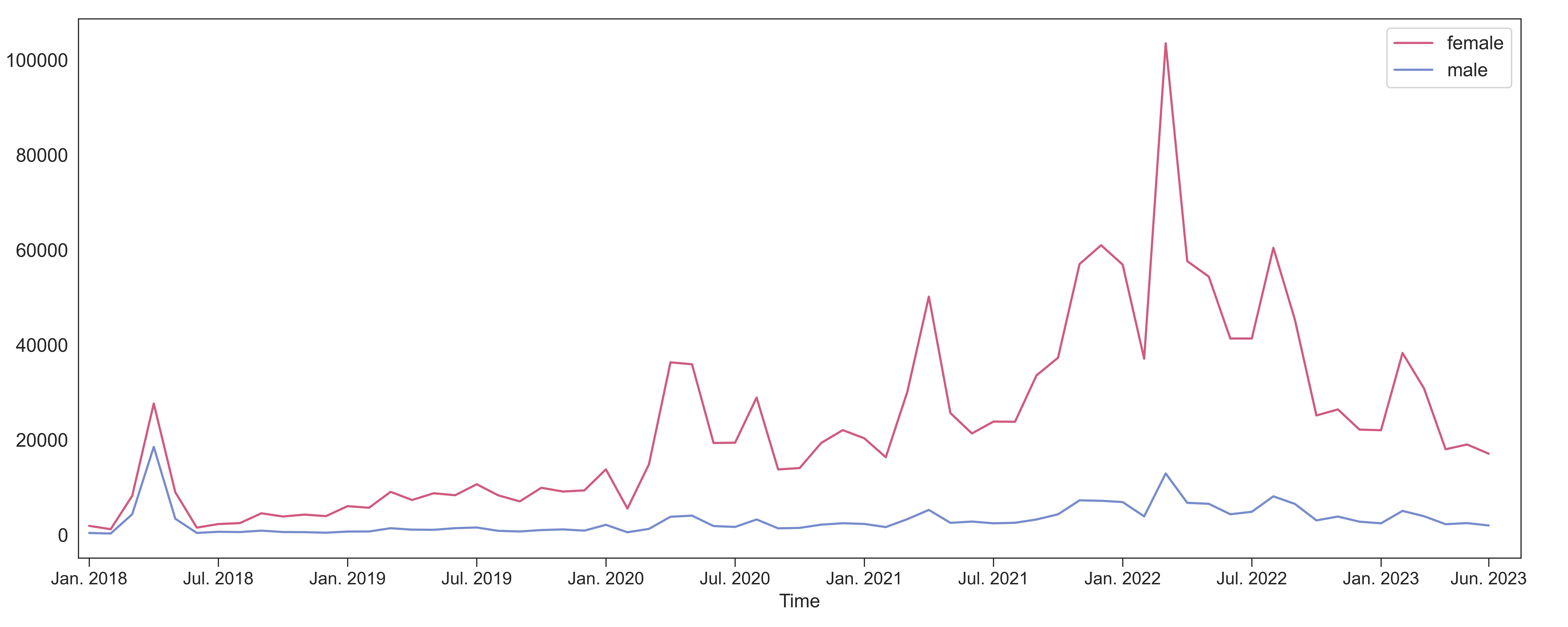

(a) Monthly number of HPV vaccine-related posts by gender

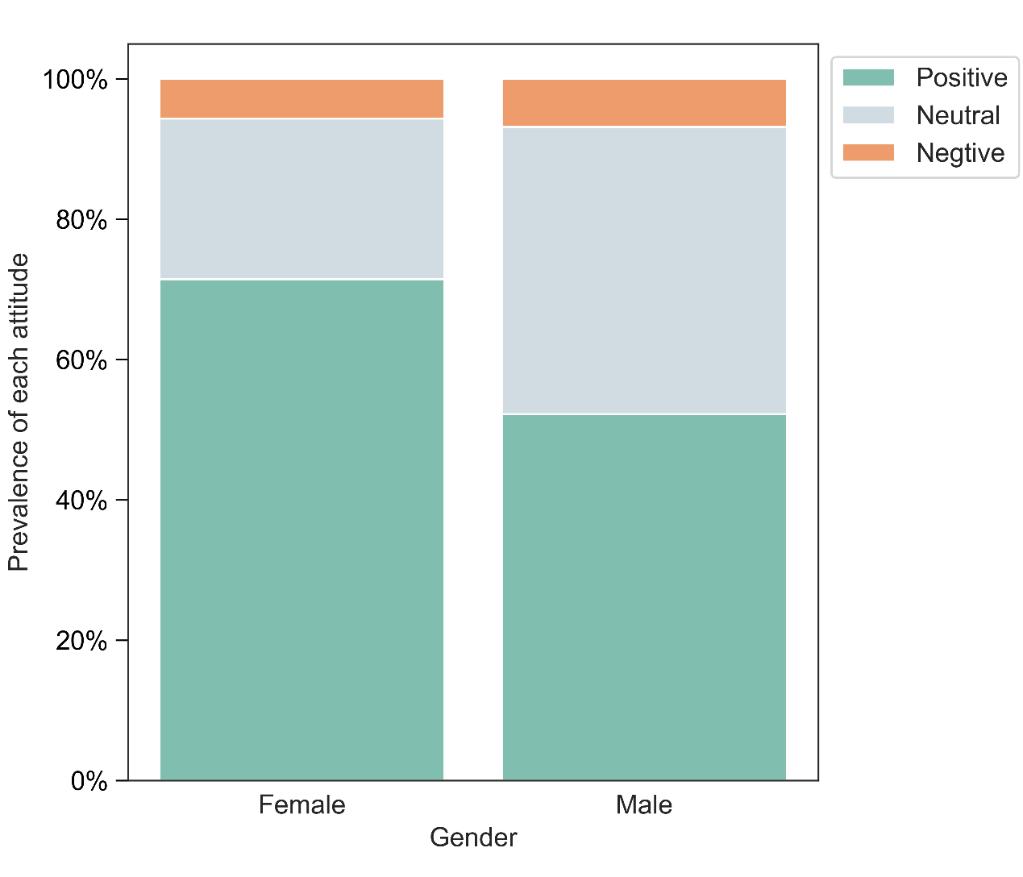

(b) Prevalence of attitudes by gender (P<.0001)

**eFigure 6.** Number of Weibo posts and prevalence of attitudes towards HPV vaccination by gender
